# Strategies for infection control and prevalence of anti-SARS-CoV-2 IgG in 4,554 employees of a university hospital in Munich, Germany

**DOI:** 10.1101/2020.10.04.20206136

**Authors:** Johanna Erber, Verena Kappler, Bernhard Haller, Hrvoje Mijočević, Ana Galhoz, Clarissa Prazeres da Costa, Friedemann Gebhardt, Natalia Graf, Dieter Hoffmann, Markus Thaler, Elke Lorenz, Hedwig Roggendorf, Florian Kohlmayer, Andreas Henkel, Michael P Menden, Jürgen Ruland, Christoph D Spinner, Ulrike Protzer, Percy Knolle, Paul Lingor, on behalf of the SeCoMRI Study Group

## Abstract

**Background:** Hospital staff are at high risk of infection during the coronavirus disease (COVID-19) pandemic. We analysed the exposure characteristics, efficacy of protective measures, and transmission dynamics in this hospital-wide prospective seroprevalence study.

**Methods and Findings:** Overall, 4554 individuals were tested for anti-severe acute respiratory syndrome coronavirus 2 (SARS-CoV-2) IgG antibodies using a chemiluminescent immunoassay. Individual risk factors, use of personal protective equipment (PPE), occupational exposure, previous SARS-CoV-2 infection, and symptoms were assessed using a questionnaire and correlated to anti-SARS-CoV-2 IgG antibody titres and PCR testing results. Odds ratios with corresponding exact 95% confidence intervals were used to evaluate associations between individual factors and seropositivity. Spatio-temporal trajectories of SARS-CoV-2-infected patients and staff mobility within the hospital were visualised to identify local hotspots of virus transmission.

The overall seroprevalence of anti-SARS-CoV-2-IgG antibody was 2.4% [95% CI 1.9–2.9]. Patient-facing staff, including those working in COVID-19 areas, had a similar probability of being seropositive as non-patient-facing staff. Prior interaction with SARS-CoV-2-infected co-workers or private contacts and unprotected exposure to COVID-19 patients increased the probability of seropositivity. Loss of smell and taste had the highest positive predictive value for seropositivity. The rate of asymptomatic SARS-CoV-2 infections was 25.9%, and higher anti-SARS-CoV-2 IgG antibody titres were observed in symptomatic individuals. Spatio-temporal hotspots of SARS-CoV-2-positive staff and patients only showed partial overlap.

**Conclusions:** Patient-facing work in a healthcare facility during the SARS-CoV-2 pandemic may be safe if adequate PPE and hygiene measures are applied. The high numbers of asymptomatic SARS-CoV-2 infections that escaped detection by symptomatic testing underline the value of cross-sectional seroprevalence studies. Unprotected contact is a major risk factor for infection and argues for the rigorous implementation of hygiene measures.

## Introduction

The outbreak of severe acute respiratory syndrome coronavirus 2 (SARS-CoV-2) was declared a pandemic and global health emergency by the World Health Organization in March 2020 and continues to pose significant challenges to healthcare systems [1]. Alarming infection rates, morbidity, and mortality have been reported among medical staff fighting the pandemic, threatening the functionality of healthcare facilities and emphasising the need to implement effective infection control measures [2-4]. The reported prevalence of anti-SARS-CoV-2 IgG antibodies in health care workers (HCW) varies from 13.7% in the New York City area, 10.2% in a nationwide study in Spain, 7.5% in 580 HCW in a Spanish hospital, 6.4% in >3000 HCW in a tertiary hospital in Belgium, 4.0% in >28790 HCW in Denmark, and 0.4% to 3.8% in Chinese hospitals [5-10]. Importantly, working in coronavirus disease (COVID-19) designated areas is reported to carry an increased risk of infection [8,11].

The greater Munich area became the epicentre of the first German SARS-CoV-2 outbreak after the first confirmed case on January 27, 2020. A rapid and massive rise in SARS-CoV-2 infections occurred in March 2020, when infected individuals returned from skiing resorts such as Ischgl, Austria, where the spread of infection was dramatic [12]. The University Hospital Munich rechts der Isar (MRI) faced the challenge of rapidly growing numbers of COVID-19 patients combined with an increasing number of staff in quarantine. To reduce the spread of infections, guidelines for the use of personal protective equipment (PPE) for staff and patients were introduced, including the obligation to wear face masks in all areas of the hospital (Fig 1A). Additionally, a telephone hotline was established to provide staff with guidance for polymerase chain reaction (PCR) testing and quarantine policies. To better understand the epidemiology and immune response to SARS-CoV-2 and to identify best-practice approaches protecting staff and patients, a prospective, observational cohort study was initiated to assess the risk factors and evidence for infection, including clinical symptoms, and to determine the seroprevalence of SARS-CoV-2 antibodies.

**Fig 1:**
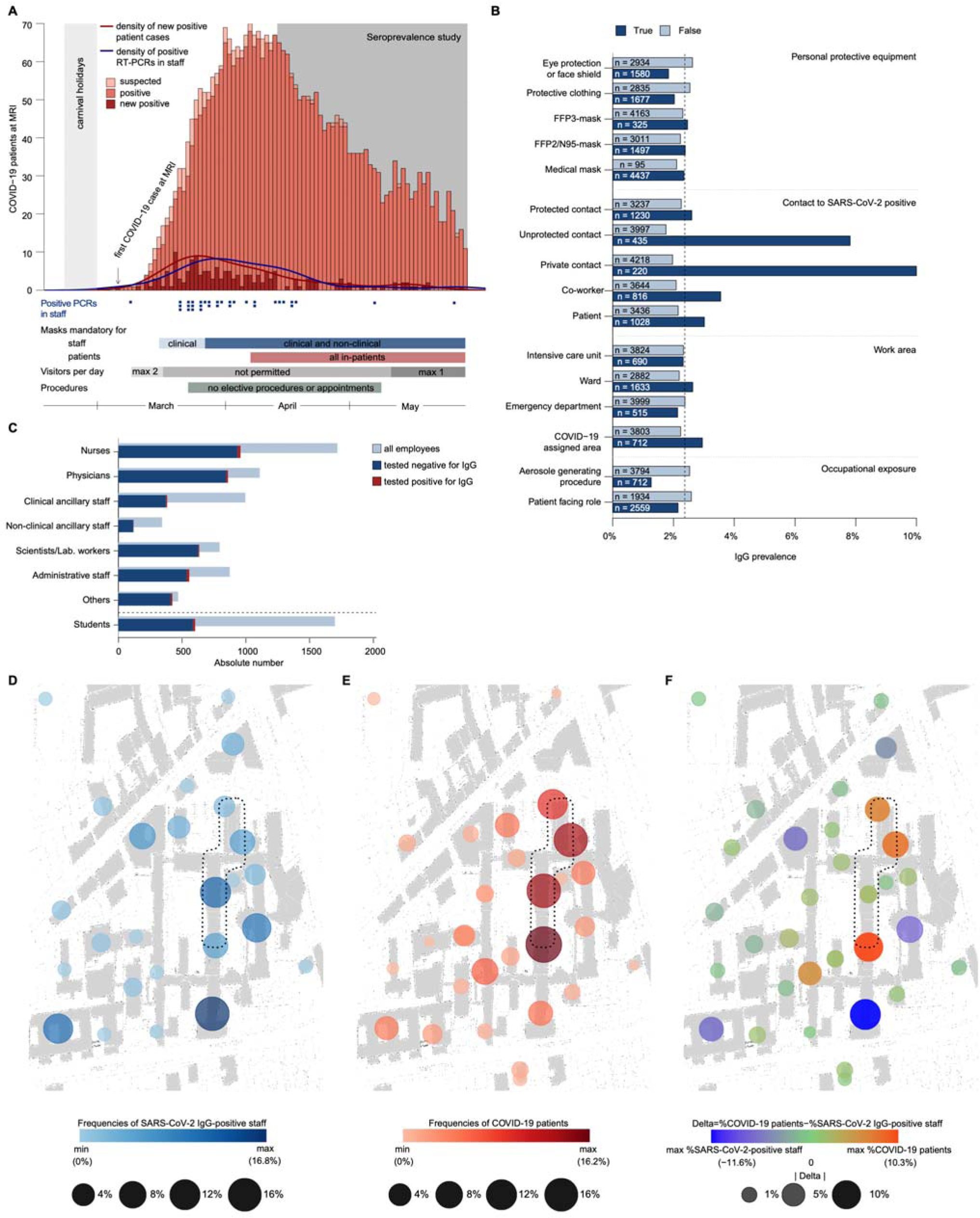
Prevalence and distribution of SARS-CoV-2 infections in patients and staff at a university hospital in Munich, Germany. **(A)** Number of all COVID-19 patients admitted to the hospital. Suspected COVID-19 cases are depicted in light orange, new confirmed COVID-19 cases are in dark red. Blue squares show positive PCR tests for SARS-CoV-2 RNA in university hospital staff. Lines indicate the densities of positive PCR tests in staff (blue) and new COVID-19 cases in patients (red). Carnival holidays: February 22, 2020 to March 1, 2020. First COVID-19 patient admitted: March 6, 2020. Seroprevalence study: April 14, 2020 to May 29, 2020. The implemented infection preventive measures are annotated below. **(B)** Prevalence of anti-SARS-CoV-2 IgG antibodies correlated to occupational risk factors. Dashed line indicates the overall seroprevalence of 2.4%. (**C**) Numbers of subjects according to occupational groups (dark blue bars, proportion of seropositive staff highlighted in red) compared to all employed staff (light blue bars). (**D–F**) Cumulative representation of frequencies of seropositive staff (**D**), COVID-19-patients (**E**), and the differences in both between different hospital areas (**F**) are shown by dot plots and assigned to distinct hospital areas. For the purposes of discretion of data from study participants, the graphical representation of spatial information is partially distorted. The dashed lines indicate COVID-19 designated areas in the hospital. FFP: Filtering face piece, Lab: Laboratory, min: Minimum, max: Maximum, MRI: University Hospital Munich rechts der Isar, PCR: Polymerase chain reaction, SARS-CoV-2: Severe acute respiratory syndrome coronavirus 2.

## Methods and Materials

### Study design and participants

Between April 14, 2020 and May 29, 2020, all clinical and non-clinical MRI staff (≥18 years) (n = 6305) and medical students of the Technical University of Munich (TUM) (n = 1699) were invited to participate in this prospective, monocentric, observational study (S1 Fig). Upon obtaining written informed consent, including the use of personal information for research, demographic data; chronic medical conditions; occupation; work location; use of PPE; exposure to SARS-CoV-2-positive patients, co-workers, or private contacts; symptom history; previous PCR testing for SARS-CoV-2, and outcome were acquired by a standardised electronic questionnaire (S2 Appendix). Serum samples were collected and subjected to anti-SARS-CoV-2 IgG and IgM testing (primary outcome). IgM was tested in all individuals until May 4, 2020 (n = 1620) and thereafter only if IgG was positive or typical symptoms of COVID-19 were reported (n = 88) (S3 Appendix, S2 Fig). Staff reporting symptoms or testing positive for IgM were recommended to undergo nasopharyngeal swab testing for SARS-CoV-2 by PCR. Personal data were stored in a pseudonymized manner using the open-source electronic case form system m4 DIS (BitCare GmbH, Munich, Germany) [13]. The study was approved by the Ethics Committee of the Technical University of Munich, School of Medicine (approval number: 216/20S).

Since March 2020, a continuous infection surveillance program for all staff has been implemented in MRI, including an employee testing centre and staff counselling (“Corona hotline”), which is available seven days per week. Individuals with symptoms compatible with COVID-19 or previous risk contacts are scheduled for testing for SARS-CoV-2 by PCR from combined nasopharyngeal swabs. Symptomatic staff or staff with high-risk contact without PPE were advised to stay home while awaiting their test results. Employees who tested negative were permitted to return to work when they had been asymptomatic for at least 48 hours. In case of a positive test result, staff were quarantined for 14 days, then retested and only allowed to continue working if the PCR tests were negative on two consecutive days. The PCR-results of staff testing were included in the present analysis if the study participants consented.

### Laboratory analysis

Primary detection of serum IgM and IgG antibodies against SARS-CoV-2 S1 or N protein was performed using a paramagnetic particle chemiluminescent immunoassay (CLIA) on an iFlash 1800 immunoassay analyser (Shenzhen Yhlo Biotech Co., Shenzhen, China). Values ≥10 AU/mL were considered positive. All sera tested positive for IgM or IgG, all sera with IgG 5–10 AU/mL, and all sera from SARS-CoV-2 PCR-positive individuals were subjected to confirmatory testing. For confirmation, total antibodies against SARS-CoV-2 N protein were determined using an electrochemiluminescent immunoassay (ECLIA) on a Cobas e411 analyser (Roche Diagnostics, Mannheim, Germany). In all samples with incongruent results, IgG antibodies against SARS-CoV-2 S1 protein were determined using an enzyme-linked immunosorbent assay (ELISA) (Euroimmun, Luebeck, Germany), and immunoblot was used to differentiate antibodies against N, S1 and the receptor binding domain of SARS-CoV-2 from those against seasonal coronaviruses (Mikrogen, Neuried, Germany) (S3 Appendix). Nucleic acids were extracted from nasopharyngeal swabs using the mSample Preparation System DNA kit following a standard protocol on an m2000sp device for RNA and DNA extraction (Abbott, Wiesbaden, Germany) and subjected to SARS-CoV-2 PCR using oligo-dT for reverse transcription and N1 and N3 primer and probe sets for detection according to the protocol of the Centre for Disease Control and Prevention, Atlanta, USA.

### Analysis of patient and staff trajectories

Anonymised patient mobility trajectory data were extracted from our hospital information system. COVID-19 was diagnosed either when patients showed typical clinical symptoms or had COVID-19-typical findings in low-dose lung CT-scan and tested positive for SARS-CoV-2 by PCR or for anti-SARS-CoV-2 IgM or IgG [14]. For spatio-temporal analysis of patient data, we used all trajectories available from December 30, 2019 to May 29, 2020 for each admitted COVID-19 patient, as the exact interval of when the patients were contagious could not be determined. Trajectories of SARS-CoV-2 IgG seropositive staff were obtained from our questionnaire data if available (February 1, 2020 to May 29, 2020). Based on the spatio-temporal trajectories of patients and staff, two types of representations were created: (i) static representations over all timeframes and (ii) dynamically animated. The static representation is based on the relative frequency of patients or staff members at each hospital location normalised by all locations of the available trajectory time. For the dynamic representation, we illustrated two different relative frequencies normalised by all timeframes: the relative frequency of individual patients in each hospital location and the relative frequency of staff members mapped to their last locations for 14 days before they tested positive for SARS-CoV-2 IgG or were quarantined.

To analyse patient mobility within the hospital during the pandemic, we compared the spatial trajectories of COVID-19 patients to all patients diagnosed with any non-COVID-19 pneumonia (viral or bacterial) from December 1, 2019 to June 10, 2020 (S3 Fig). All analyses were performed using R software version 3.6.0 (R Foundation for Statistical Computing, Vienna, Austria), and the source code is available at https://github.com/AnaGalhoz37/SeCOMRI.

### Statistical analysis

Absolute and relative frequencies of positive tests for anti-SARS-CoV-2 IgG and IgM (CLIA, Yhlo) are given for all study participants and relevant subgroups. An exact 95% confidence interval (CI) for the estimated seroprevalence is presented. To evaluate the association with potential risk factors, odds ratios (ORs) with corresponding exact 95% CIs (mid-p intervals) were estimated. The distributions of antibody titres are visualised by boxplots or dot plots and are described by medians and quartiles. Spearman’ s rank correlation coefficient was used to evaluate the association between the time of IgG testing and the IgG titre level. The confidence interval widths were not adjusted for multiplicity. Missing data were not imputed, and the number of missing values is presented for each variable. Statistical analyses were conducted using the statistical software R version 4.0.2 (R Foundation for Statistical Computing, Vienna, Austria).

## Results

### Anti-SARS-CoV-2-IgG seroprevalence in 4,554 hospital employees

The study participation rate was 63.5% [4001/6305] for employees and 35.5% [603/1699] for medical students; the complete data from 4554 individuals were available for primary analysis (S1 Fig). The mean age of the study participants was 38.5 years; 3207 (70.4%) were women and 1342 (29.5%) were men (S4 Fig). Positive anti-SARS-CoV-2 IgG antibodies were found in 108/4554 study participants. In 102 individuals, additional assays confirmed the anti-SARS-CoV-2 IgG screening result (S1 Table). Two additional individuals reporting a positive PCR test seroconverted during follow-up. Four individuals with IgG titres of 5–10 AU/mL in the screening assay, which is below the cut-off, were found to be positive in at least two other assays (S1 and S2 Tables). In five individuals, the screening result could not be confirmed by the other assays used; in one, there was insufficient material available to complete testing (S3 Table) (see S3 Appendix for details). Considering all 108 study participants who tested positive for anti-SARS-CoV-2 IgG antibodies in at least two different assays, we determined a seroprevalence of 2.4% [95% CI, 1.9–2.9] (primary endpoint).

### Individual and occupational risk factors for SARS-CoV-2 infection

The first patient with PCR-confirmed COVID-19 was admitted to our university hospital on March 6, 2020, and 163 COVID-19 patients were hospitalised between March 6, 2020 and May 29, 2020 (Fig 1A). Infection prevention measures were dynamically adjusted according to the prevalent pandemic situation (Fig 1A). Individual risk factors for infection were identified through correlation of self-reported survey data with seropositivity for anti-SARS-CoV-2 IgG antibodies. We found an association between seropositivity and male sex (OR 1.54 [95% CI, 1.03–2.27]) or age, with the highest frequency observed for the age group of 51–60 years (OR 1.75 [95% CI, 1.06–2.85] compared to those ≤30 years) (Table 1, 4, Fig 5). A higher relative frequency of seropositivity was found for individuals with diabetes mellitus (OR 2.96 [95% CI, 1.01–6.81], while no significant differences were observed in staff with pre-existing pulmonary or cardiovascular disease (Table 2, S5 Fig). Seropositivity was decreased in smokers (OR 0.52 [95% CI, 0.26–0.94]) (Table 2, S5 Fig). No relevant difference in seropositivity was observed between HCW involved in direct patient care, including care of COVID-19 patients, and those working in intensive care units or the emergency department compared to staff members not working in these areas and not performing patient-associated tasks (Fig 1B, Table 2, S5 Fig). Conversely, seropositivity was particularly high in administrative staff without any direct patient contact (OR 2.36 [95% CI, 1.19–4.80]) (Fig 1C, Table 1, S5 Fig). Seropositivity was also markedly increased in staff who declared exposure to co-workers (OR 1.74 [95% CI, 1.11–2.65]) or private contacts with SARS-CoV-2 infection (OR 5.56 [95% CI, 3.32–8.94]) (Fig 1B, Table 2, S5 Fig). Self-reported unprotected contact with COVID-19 patients (no medical mask, <1.5 metre distance, or aerosol-generating medical procedure without filtering masks with either FFP2 or N95 standard and/or eye protection or face shields) was associated with higher seroprevalence (OR 4.77 [95% CI, 3.09–7.22]) (Table 2, S5 Fig). For staff reporting to perform aerosol-generating procedures we observed an even lower rate of seropositivity (OR 0.50 [95% CI 0.23-0.94]; Fig 1B, Table 2, S5 Fig).

**Table 1:**
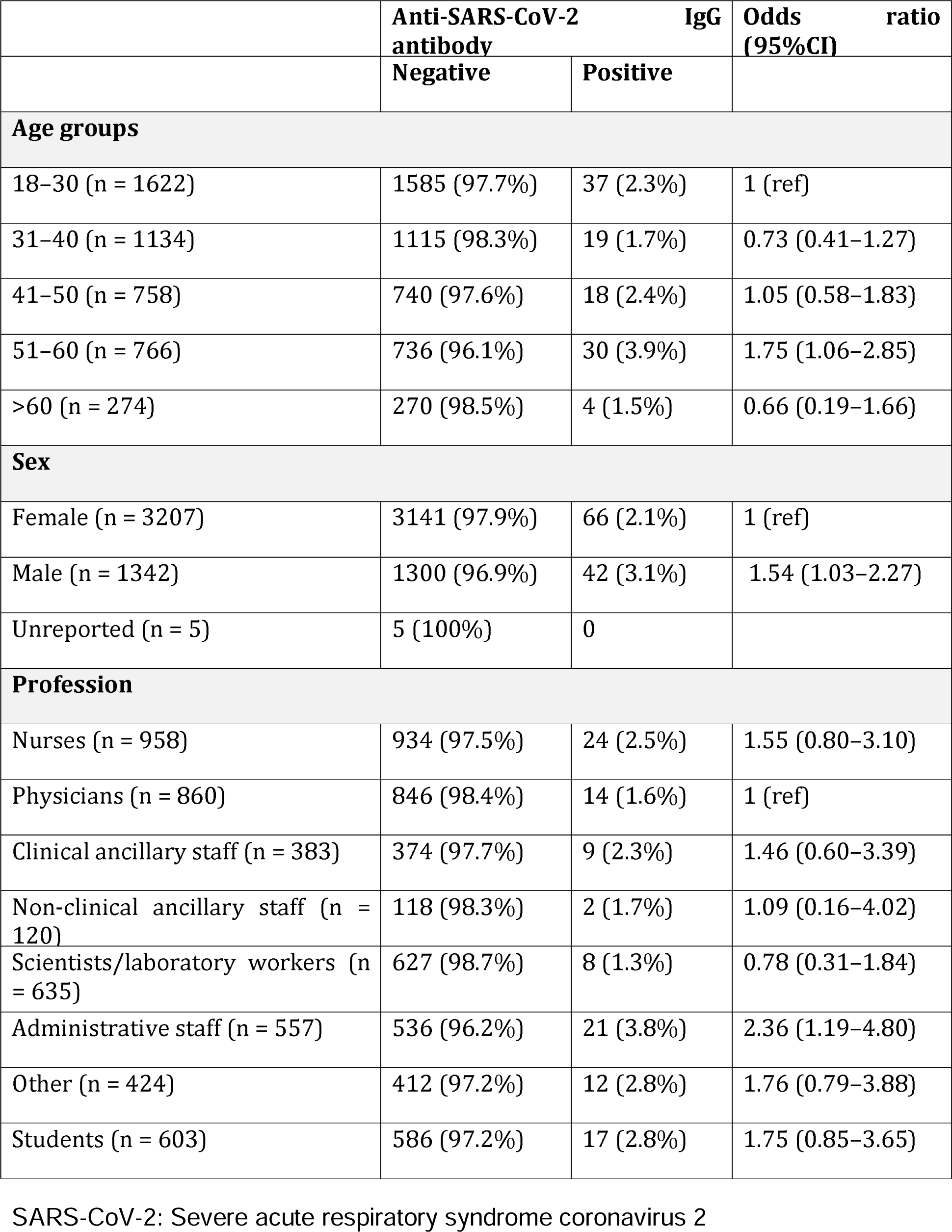
Seroprevalence by general characteristics and occupation.

**Table 2:**
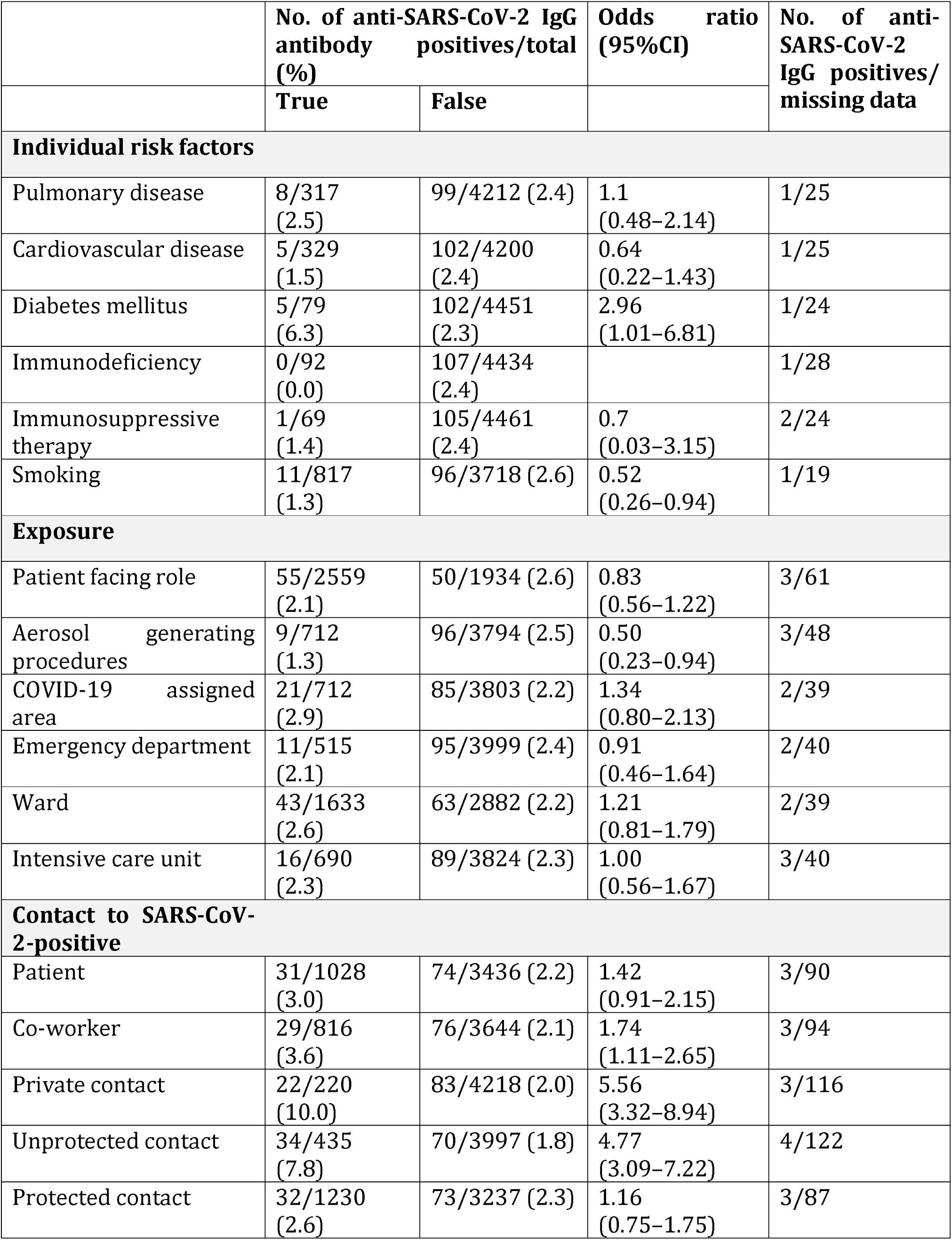
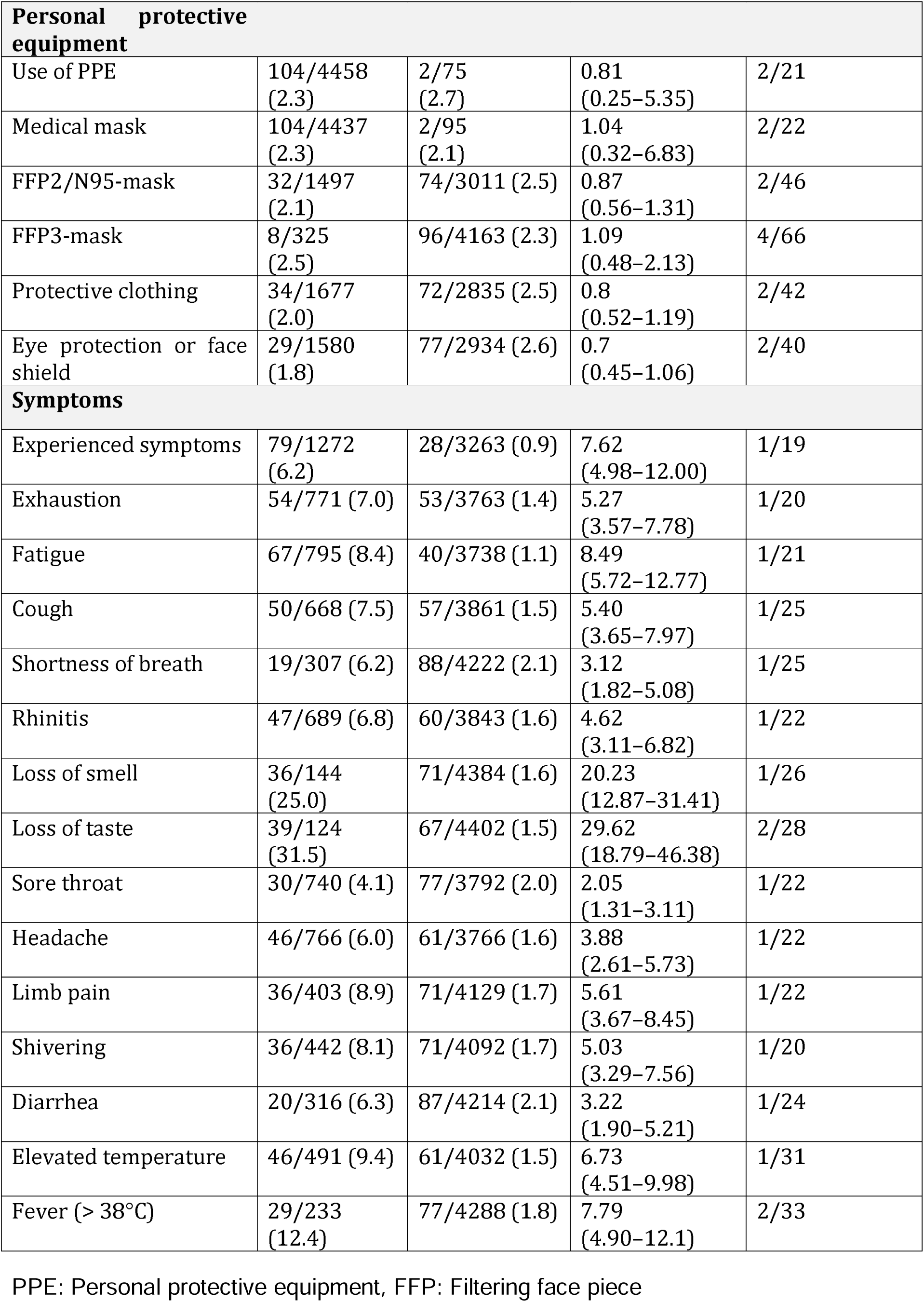
Seroprevalence by self-reported risk factors and symptoms.

|  | No. of anti-SARS-CoV-2 IgG antibody positives/total (%) |  | Odds ratio (95%CI) | No. of anti-SARS-CoV-2 IgG positives/missing data |
| --- | --- | --- | --- | --- |
|  | True | False |  |  |
| Individual risk factors |  |  |  |  |
| Pulmonary disease | 8/317 (2.5) | 99/4212 (2.4) | 1.1 (0.48–2.14) | 1/25 |
| Cardiovascular disease | 5/329 (1.5) | 102/4200 (2.4) | 0.64 (0.22–1.43) | 1/25 |
| Diabetes mellitus | 5/79 (6.3) | 102/4451 (2.3) | 2.96 (1.01–6.81) | 1/24 |
| Immunodeficiency | 0/92 (0.0) | 107/4434 (2.4) |  | 1/28 |
| Immunosuppressive therapy | 1/69 (1.4) | 105/4461 (2.4) | 0.7 (0.03–3.15) | 2/24 |
| Smoking | 11/817 (1.3) | 96/3718 (2.6) | 0.52 (0.26–0.94) | 1/19 |
| Exposure |  |  |  |  |
| Patient facing role | 55/2559 (2.1) | 50/1934 (2.6) | 0.83 (0.56–1.22) | 3/61 |
| Aerosol generating procedures | 9/712 (1.3) | 96/3794 (2.5) | 0.50 (0.23–0.94) | 3/48 |
| COVID-19 assigned area | 21/712 (2.9) | 85/3803 (2.2) | 1.34 (0.80–2.13) | 2/39 |
| Emergency department | 11/515 (2.1) | 95/3999 (2.4) | 0.91 (0.46–1.64) | 2/40 |
| Ward | 43/1633 (2.6) | 63/2882 (2.2) | 1.21 (0.81–1.79) | 2/39 |
| Intensive care unit | 16/690 (2.3) | 89/3824 (2.3) | 1.00 (0.56–1.67) | 3/40 |
| Contact to SARS-CoV-2-positive |  |  |  |  |
| Patient | 31/1028 (3.0) | 74/3436 (2.2) | 1.42 (0.91–2.15) | 3/90 |
| Co-worker | 29/816 (3.6) | 76/3644 (2.1) | 1.74 (1.11–2.65) | 3/94 |
| Private contact | 22/220 (10.0) | 83/4218 (2.0) | 5.56 (3.32–8.94) | 3/116 |
| Unprotected contact | 34/435 (7.8) | 70/3997 (1.8) | 4.77 (3.09–7.22) | 4/122 |
| Protected contact | 32/1230 (2.6) | 73/3237 (2.3) | 1.16 (0.75–1.75) | 3/87 |

| **Personal protective equipment** |  |  |  |  |
| --- | --- | --- | --- | --- |
| Use of PPE | 104/4458<br>(2.3) | 2/75<br>(2.7) | 0.81<br>(0.25–5.35) | 2/21 |
| Medical mask | 104/4437<br>(2.3) | 2/95<br>(2.1) | 1.04<br>(0.32–6.83) | 2/22 |
| FFP2/N95-mask | 32/1497<br>(2.1) | 74/3011 (2.5) | 0.87<br>(0.56–1.31) | 2/46 |
| FFP3-mask | 8/325<br>(2.5) | 96/4163 (2.3) | 1.09<br>(0.48–2.13) | 4/66 |
| Protective clothing | 34/1677<br>(2.0) | 72/2835 (2.5) | 0.8<br>(0.52–1.19) | 2/42 |
| Eye protection or face shield | 29/1580<br>(1.8) | 77/2934 (2.6) | 0.7<br>(0.45–1.06) | 2/40 |
| **Symptoms** |  |  |  |  |
| Experienced symptoms | 79/1272<br>(6.2) | 28/3263 (0.9) | 7.62<br>(4.98–12.00) | 1/19 |
| Exhaustion | 54/771 (7.0) | 53/3763 (1.4) | 5.27<br>(3.57–7.78) | 1/20 |
| Fatigue | 67/795 (8.4) | 40/3738 (1.1) | 8.49<br>(5.72–12.77) | 1/21 |
| Cough | 50/668 (7.5) | 57/3861 (1.5) | 5.40<br>(3.65–7.97) | 1/25 |
| Shortness of breath | 19/307 (6.2) | 88/4222 (2.1) | 3.12<br>(1.82–5.08) | 1/25 |
| Rhinitis | 47/689 (6.8) | 60/3843 (1.6) | 4.62<br>(3.11–6.82) | 1/22 |
| Loss of smell | 36/144<br>(25.0) | 71/4384 (1.6) | 20.23<br>(12.87–31.41) | 1/26 |
| Loss of taste | 39/124<br>(31.5) | 67/4402 (1.5) | 29.62<br>(18.79–46.38) | 2/28 |
| Sore throat | 30/740 (4.1) | 77/3792 (2.0) | 2.05<br>(1.31–3.11) | 1/22 |
| Headache | 46/766 (6.0) | 61/3766 (1.6) | 3.88<br>(2.61–5.73) | 1/22 |
| Limb pain | 36/403 (8.9) | 71/4129 (1.7) | 5.61<br>(3.67–8.45) | 1/22 |
| Shivering | 36/442 (8.1) | 71/4092 (1.7) | 5.03<br>(3.29–7.56) | 1/20 |
| Diarrhea | 20/316 (6.3) | 87/4214 (2.1) | 3.22<br>(1.90–5.21) | 1/24 |
| Elevated temperature | 46/491 (9.4) | 61/4032 (1.5) | 6.73<br>(4.51–9.98) | 1/31 |
| Fever (> 38°C) | 29/233<br>(12.4) | 77/4288 (1.8) | 7.79<br>(4.90–12.1) | 2/33 |

PPE: Personal protective equipment, FFP: Filtering face piece

### Symptoms and anti-SARS-CoV-2 IgG antibody titres in hospital staff

In our cohort, 1272 individuals (27·9%) reported current or recent (within eight weeks before testing) presence of at least one symptom indicative of COVID-19 (Table 2, S5 Fig), 79 of which (6.2%) were seropositive for anti-SARS-CoV-2 IgG antibodies (Table 2, S5 Fig). Loss of smell (36 seropositive of 144 individuals with reported loss of smell, 25.0%) and loss of taste (39 of 124, 31.5%) had the highest positive predictive value (Table 2, S5 Fig), and seropositivity was associated with a higher number of symptoms reported (Table 3, S5 Fig).

**Table 3:** Seroprevalence by symptom onset and frequency.

|  | SARS-CoV-2 IgG |  | Odds ratio (95%CI) |
| --- | --- | --- | --- |
|  | Negative | Positive |  |
| **Symptom onset** |  |  |  |
| Not applicable (n = 3373) | 3336 (98.9%) | 37 (1.1%) | 1 (ref) |
| Past 14 days (n = 219) | 209 (95.4%) | 10 (4.6%) | 4.36 (2.02–8.59) |
| Past 3 to 8 weeks (n = 943) | 883 (93.6%) | 60 (6.4%) | 6.113 (4.05–9.35) |
| Unknown (n = 19) | 18 | 1 |  |
| **Symptom frequency (p < 0.001)** |  |  |  |
| 0 (n = 3273) | 3245 (99.1%) | 28 (0.9%) | 1 (ref) |
| 1–4 (n = 548) | 529 (96.5%) | 19 (3.5%) | 4.17 (2.27–7.5) |
| 5–8 (n = 491) | 454 (92.5%) | 37 (7.5%) | 9.42 (5.72–15.7) |
| 9–14 (n = 223) | 200 (89.7%) | 23 (10.3%) | 13.32 (7.45–23.58) |
| Unknown (n = 19) | 18 | 1 |  |

In seropositive individuals, we found no significant differences in anti-SARS-CoV-2 IgG antibody titres for different age groups, sex, comorbidities, or exposure profiles (S6 Fig). However, anti-SARS-CoV-2 IgG levels were higher in staff who reported more COVID-19-related symptoms (S7 Fig). We observed the highest titres for those who reported diarrhoea, fever, elevated temperature, shivering, limb pain, and headache (S7 Fig).

### Value of symptom-based PCR testing

Symptom-based PCR testing of material obtained from nasopharyngeal swabs was initiated early during the pandemic through a dedicated COVID-19 telephone hotline. The first hospital employee with SARS-CoV-2 infection was identified on March 9, 2020, and 28 individuals with SARS-CoV-2 infection detected by PCR before participating in this study tested positive for anti-SARS-CoV-2 IgG (Fig 1A), (S8 Fig). Ten seropositive individuals reported positive PCR, one positive antibody testing at other facilities. However, 68 of 108 (63%) SARS-CoV-2 infections had not been diagnosed previously, data on prior testing was missing for one individual. Among these 68 seropositive individuals, 28 did not report any COVID-19-typical symptoms in the initial survey (25.9% of all seropositive staff), indicating that symptom-based testing can miss SARS-CoV-2 infection.

### Analysis of spatio-temporal trajectories

To identify and localize potential hotspots of infection, we systematically analysed contact between staff and COVID-19 patients using the cumulative data on staff serology and the patient registry. Thus, we plotted available spatial and temporal information on the presence of COVID-19 patients and anti-SARS-CoV-2 IgG antibody-positive staff with daily resolution on a hospital map. Visualisation of these spatio-temporal mobility trajectories revealed only a slight overlap between the distinct spatial and temporal hotspots of COVID-19 patients and anti-SARS-CoV-2 IgG antibody-positive staff (Fig 1D, Videos).

## Discussion

Studies on the seroprevalence of SARS-CoV-2 are essential to reliably analyse exposure characteristics among hospital staff and to evaluate the efficacy of protective measures. Despite the high overall burden of patients with COVID-19 disease in our hospital, the seroprevalence of 2.4% for anti-SARS-CoV-2 IgG antibodies among university hospital staff after the first wave in Germany is lower than that reported in previous studies, which may be attributed to differences in the cohort composition, fast implementation of protective measures, or frequency of exposure [1,2].

Although hospital staff have an increased occupational risk of contact with SARS-CoV-2-infected patients, and a high burden of SARS-CoV-2 infection among HCW involved in the care of COVID-19 patients has been previously reported, we did not observe higher seroprevalences in staff who reported direct patient contact, including those working in COVID-19 designated areas [11,12,15]. Unexpectedly, we observed lower seroprevalence in staff who reported exposure to aerosol-generating high-risk procedures, possibly reflecting increased awareness and the implementation of particularly rigorous personal protection measures in this subgroup. Furthermore, the type of protective equipment used was not associated with seroprevalence, but 98% of staff reported routinely using medical masks, which was required by internal hospital policy for patient-facing staff since March 16, 2020, and for all staff starting March 27, 2020. The almost parallel rise of SARS-CoV-2 infection cases in staff and patients is suggestive of extrinsic infection causes in both groups, such as the simultaneous return from high-risk holiday areas. Consistently, exposure to SARS-CoV-2-infected private contacts or co-workers was the most important risk factor for SARS-CoV-2 infection in our cohort. This underscores the need for adherence to protective measures during both private and professional personal contacts.

Male staff in our study cohort showed a higher seroprevalence. Based on a recent study reporting a significantly lower perceived infection risk in men than in women, adherence to hygiene guidelines and social distancing measures might have been lower in male staff [16,17]. Interestingly, smokers showed a significantly lower seroprevalence, which is in contrast to that reported in previous studies [6,7,9,10,16]. Because smokers are more susceptible to respiratory tract infections and smoking involves hand-to-mouth contact and frequent social interactions, the lower seroprevalence is unexpected but in line with other reports [18,19,20-22]. Staff suffering from diabetes mellitus had a higher seroprevalence than non-diabetics. Previously, diabetes mellitus reportedly correlates with the severity of COVID-19 and associated mortality, but no increase in susceptibility to SARS-CoV-2 infection have been reported to date [23].

The serological assessment confirmed infection in most individuals who reported positive testing for SARS-CoV-2 by PCR. However, repeated testing over >4 weeks with at least two separate assays each did not detect antibodies in six individuals (two with positive in-house PCR results and four with reported positive external PCR results) (S8 Fig, No. 14, 29). This may be explained by false-positive PCR results or by the failure to develop antibody responses after low-symptomatic infection, which may occur in up to 10% of convalescents following SARS-CoV-2 infection [24].

For 19 seropositive individuals, previous PCR testing yielded negative results. Because virus shedding is often limited to only a few days, PCR detection of SARS-CoV-2 RNA critically depends on timing and properly performed nasopharyngeal swabs. Thus, negative PCR test results are frequent and should be questioned, repeated, and complemented by antibody testing if clinical signs or history suggest an infection [25]. However, false-positive antibody test results also need to be considered, particularly in the light of an overall low prevalence and possible cross-reactivity with other coronaviruses. The IgG immunoassay used for screening proved to have a specificity of 99.89% in our study; it uses two SARS-CoV-2 antigens (N and S1) for detection and has an estimated sensitivity of 96.30%. We retested all IgM or IgG-positive sera and all sera with titres of 5–10 AU/mL, which is below the cut-off, by a second assay with particularly high specificity (99.90%) that uses recombinant N-protein as the antigen. If required, additional assays were performed: either an ELISA, using recombinant S1 protein as capture antigen, or an immunoblot, testing for antibodies against three different SARS-CoV-2 antigens. Serum was only considered positive if two or more antibody assays were positive (S3 Appendix). However, the requirement of such extensive confirmatory testing strengthens the notion that each test for SARS-CoV-2 antibodies requires critical evaluation.

Our study also revealed that approximately 2/3 of the seropositive staff had a hitherto undetected infection. These infections may have been oligo- or asymptomatic without even alarming medically trained personnel. Furthermore, 25 anti-SARS-CoV-2 IgG antibody-positive individuals had not been PCR-tested despite reporting at least one COVID-19 compatible symptom. 1183 staff members tested seronegative despite reporting at least one symptom related to COVID-19. The focus on symptoms with the strongest association with seropositivity, such as loss of smell, loss of taste, fatigue, fever, and cough, may therefore be helpful in developing more accurate and economical screening algorithms. Our results highlight that symptom-based testing may miss staff infections. All 28 asymptomatic seropositive individuals remained undiagnosed before the study, emphasising the significance of rigorous implementation of systematic hygiene measures in pandemic situations. Transmission by asymptomatic infected staff may occur at any time and will not be prevented by random testing. All these results strongly support the continuous use of medical masks as a simple and efficient measure for employee and patient protection.

To identify infection hotspots and putative patient/staff overlaps, we visualised the temporo-spatial mobility trajectories of patients and staff to monitor the infection dynamics. Real-time use of such trajectory mapping at high resolution may yield additional information that allows the reaction to react more quickly and intuitively to infection foci. Continuous evaluation of mobility trajectory mappings may highlight areas of recurrent infections and thus identify previously unattended needs that should be addressed for future waves of the pandemic.

The current study has several limitations. First, because this was a voluntary assessment, participation was incomplete and may have biased the results. We cannot exclude the possibility that staff members with a higher perceived risk of infection were more likely to participate. Furthermore, as most students stayed at home during lock-down, their participation rate was low. Second, symptoms and exposures were retrospectively assessed and self-reported and thus subject to a recall bias. Third, although we attempted a cross-sectional analysis, our data documents the average seroprevalence during the entire testing period; thus, seroconversions occurring during this period may have been missed. Finally, PCR testing results were only available from individuals who consented to their use (4373/4554), limiting the possibility of cross-validating PCR results with seroprevalence.

Altogether, our findings have several important implications. We did not observe a relevant increase in anti-SARS-CoV-2 IgG antibody seropositivity in HCW (including those working with COVID-19 patients) compared to staff who were not directly involved in patient care as long as PPE was used. Importantly, interaction with SARS-CoV-2 infected co-workers and/or private contacts was a major risk factor for infection. Among the symptoms reported, loss of smell and taste were strongly indicative of SARS-CoV-2 infection. However, almost two-thirds of seropositive individuals detected in this cross-sectional study had not been previously diagnosed with SARS-CoV-2 infection, while 25.9% reported asymptomatic infections. Thus, obligatory wearing of medical masks by all employees and, whenever tolerated, also by patients, may minimise virus transmission risks, although it was not possible to formally separate this effect from that of minimising personal contacts imposed by the general lock-down. To date, the value of anti-SARS-CoV-2 antibodies for protective immunity and their sustainability in infected individuals remains unclear. Longitudinal studies with combined testing for virus-specific antibodies and their infection-neutralizing ability, as well as virus-specific T-cell immunity are needed to estimate the longevity and protective value of the anti-SARS-CoV-2 IgG antibody responses in hospital staff.

## Data Availability

Anonymised aggregated data will be available to researchers upon reasonable request, which should be directed to the corresponding author. The source code of the trajectory analysis is available at https://github.com/AnaGalhoz37/SeCOMRI.

## Abbreviations

CI: confidence interval
CLIA: chemiluminescent immunoassay
ECLIA: electrochemiluminescent immunoassay
ELISA: enzyme-linked immunosorbent assay
HCW: health care workers
MRI: University Hospital Munich rechts der Isar
OR: odds ratio
PCR: polymerase chain reaction
PPE: personal protective equipment
SARS-CoV-2: severe acute respiratory syndrome coronavirus 2

## Acknowledgements

A list of the SeCoMRI Study Group is available in S1 Appendix in the Supporting Information. We are indebted to all study participants who consented to having their data published. This study would not have been possible without the contribution of clinical and administrative staff involved in the enrolment and obtaining informed consent of study participants. In particular, we would like to thank all technical staff of the Institute of Virology and the Institute of Clinical Chemistry for performing the diagnostic tests. We are grateful to Yhlo, Shenzhen, China, and Mikrogen, Neuried, Germany, for providing part of the test kits for free. We are obliged to the hospital board of directors for their financial and organisational support.

## Supporting Information files

**S1 Appendix: SeCoMRI Study Group**

**S2 Appendix: Questionnaire**

**S3 Appendix: Calculation of specificity and sensitivity of the SARS-CoV-2 antibody tests**

**S1 Table: Samples with confirmed positive IgG against SARS-CoV-2**

**S2 Table: Summary of confirmatory assays**

**S3 Table: Samples with positive IgG against SARS-CoV-2 that could not be confirmed**

**S4 Table: Samples with positive IgM against SARS-CoV-2 that could not be confirmed**

**S5 Table: Calculation of specificity and sensitivity**

**S1 Figure: Study flow chart illustrating the testing algorithm and included results**

**S2 Figure: Anti-SARS-CoV-2 IgG and IgM levels**

**S3 Figure: Relative frequency of requested diagnostics, therapies, and spatial information between patients diagnosed with COVID-19 and non-COVID-19 pneumonia**

**S4 Figure: Age and sex distribution of the study participants**

**S5 Figure: Graphic representation of all odds ratios for seropositivity to SARS- CoV-2 IgG represented in Table S1–3**

**S6 Figure: Distribution of anti-SARS-CoV-2 IgG levels stratified for personal/occupational risk factors**

**S7 Figure: Anti-SARS-CoV-2 IgG levels and symptoms**

**S8 Figure: Time course of anti-SARS-CoV-2 antibody and PCR test results in 33 employees**

**S1 Video: Time lapse of the relative frequencies of all trajectories for COVID-19 cases**

**S2 Video: Time lapse of the relative frequencies of trajectories available for SARS-CoV-2-positive staff working on campus**

**S3 Video: Time lapse of the relative frequencies for the difference in trajectories available for COVID-19 cases and for SARS-CoV-2-positive staff**

