## Supplemental Material for "Strategies for infection control and prevalence of anti-SARS-CoV-2 IgG in 4,554 employees of a university hospital in Munich, Germany"

**to**

S1 Appendix: SeCoMRI Study Group

Balqees Al Darweesh

Clara Balzer

Felix Bauerdorf

Alexander Böhner

Dirk Busch

Lisena Cala

Ana Cirac
Adam Chaker

Anaïs Marie Theresa Doll

Johanna Erber

Manon Feuchtinger

Ana Galhoz
Friedemann, Gebhardt

Marisa Geisberger
Markus Gerhard

Oliver Goldhardt

Katharina Gresset-Kaliebe

Natalia Graf

Florian Groß

Roman Günthner

Martin Halle

Bernhard Haller

Joachim Hellemann

Andreas Henkel

Maximilian Hinz

Dieter Hoffmann
Klaus-Peter Janssen

Robert Kaczmarczyk

Verena Kappler

Percy Knolle

Florian Kohlmayer

Susanne Kossatz

Klaus Kuhn

Zsuzsanna Kurgyis

Vincent Lallinger

Judith Lammer

Paul Lingor
Elke Lorenz

Felix Mayr

Michael M. Menden
Hrvoje Mijočević

Caroline Sandra Moesta

Ruth Neuhauser

Andrea Pagani

Anna Caroline Pilz
Clarissa Prazeres da Costa

Sarah Preis
Ulrike Protzer
Michael Quante
Hedwig Roggendorf
Jürgen Ruland

Mine Sargut

Cora Scheerer

Roland M. Schmid

Paul Schmidle

Christine Schönmann

Florian Schraml
Christoph D. Spinner

Annette Susanne Steimle-Grauer

Christian Stöß

Pavel Stupakov

Markus Thaler

Dolores Thum

Dirk Tomsitz
Wolfgang Weber
Angelika Werner
Christof Winter

S2 Appendix: Questionnaire

The original survey was in German and was acquired by a standardized, electronic questionnaire.

Baseline characteristics

Date of assessment	□□.□□.□□□□

Baseline characteristic: Age, sex

Department:

Worksite:

Occupation

□ Physician □ Nurse

□ Lab Worker □ Hygiene Staff □ Clinical Ancillary Staff □ Cleaning Staff □ Patient Transport

□ Administration □ Technical Staff □ IT □ Scientist □Student □ Others

Do you have a patient facing role?	□ Yes	□ No

Exposure and personal protective equipment

|  | In which area(s) have you been placed? (multiple answers possible) |
| --- | --- |

| Work area | Currently | Past 48 hours | Past 3 to 14 days | Past 3 to 8 weeks |
| --- | --- | --- | --- | --- |
| COVID-19 assigned area |  |  |  |  |
| Emergency department |  |  |  |  |
| Ward |  |  |  |  |
| Intensive care unit |  |  |  |  |
| Other |  |  |  |  |
| Aerosol generating procedures* |  |  |  |  |

*Endoscopy, bronchoscopy, tracheal intubation, non-invasive ventilation, transoesophageal echo, etc.

Have you been in contact with SARS-CoV-2-positive individuals? □ Yes	□ No

| COVID-19 contact | Currently | Past 48 hours | Past 3 to 14 days | Past 3 to 8 weeks |
| --- | --- | --- | --- | --- |
| Patients at MRI |  |  |  |  |
| Co-worker at MRI |  |  |  |  |
| Private contact |  |  |  |  |
| Protected (mask and physical distance, or FFP2/N95 and eye protection when performing aerosol-generating procedures |  |  |  |  |
| Unprotected (none of the abovementioned, or mask only when performing aerosol generating procedures |  |  |  |  |

Do you use personal protective equipment?	□ Yes	□ No

|  | If so, which ones? (multiple answers possible) |
| --- | --- |

| Personal protective equipment | Currently | Past 48 hours | Past 3 to 14 days | Past 3 to 8 weeks |
| --- | --- | --- | --- | --- |
| Mask |  |  |  |  |
| FFP2/N95 |  |  |  |  |
| FFP3 |  |  |  |  |
| Protective clothing |  |  |  |  |
| Eye protection or face shield |  |  |  |  |
| Others |  |  |  |  |

Individual factors

What applies to you (multiple answers possible)?

□ Smoking □ Pulmonary disease □ Cardiovascular disease □ Diabetes mellitus

□ Immunodeficiency	□ Immunosuppressive therapy

□ Other

Have you had COVID-19 compatible symptoms? □ Yes	□ No (multiple answers possible)

| Symptoms | Currently | Past 48 hours | Past 3 to 14 days | Past 3 to 8 weeks |
| --- | --- | --- | --- | --- |
| Exhaustion |  |  |  |  |
| Fatigue |  |  |  |  |
| Cough |  |  |  |  |
| Shortness of breath |  |  |  |  |
| Rhinitis |  |  |  |  |
| Loss of smell |  |  |  |  |
| Loss of taste |  |  |  |  |
| Sore throat |  |  |  |  |
| Headache |  |  |  |  |
| Limb pain |  |  |  |  |
| Shivering |  |  |  |  |
| Diarrhoea |  |  |  |  |
| Elevated temperature (37.3–37.9°C) |  |  |  |  |
| Fever (> 38°C) |  |  |  |  |
| Current body temperature □□ . □□ °C | | | | |

Have you ever been tested for SARS-CoV-2?	□ Yes	□ No

if so:	□	Past 14 days □	More than 14 days ago

Where? □ MRI	□ Registered physician	□ Department Of Public Order

How? □ Nasopharyngeal swab	□ Blood	□	Stool

Test result:

□ Pending	□ Positive for SARS-COV-2 □ Negative for SARS-CoV-2

COVID-19 disease

Have you already had COVID-19?	□ Yes	□ No

Treatment:	□ Outpatient/at home □ Inpatient/normal ward □ Ward and intensive care unit

S3 Appendix: Calculation of specificity and sensitivity of the SARS-CoV-2 antibody tests

IgG and IgM antibodies were determined in 4554 and 1708 sera, respectively, using a paramagnetic particle chemiluminescent immunoassay (CLIA) on an iFlash 1800 immunoassay analyser (Shenzhen Yhlo Biotech Co., Shenzhen, China). This assay was selected as a screening assay as it detects antibodies directed against either SARS-CoV-2 S1 or N protein. According to the manufacturer’s instructions, values ≥ 10 AU/mL were considered positive. Anti-SARS-CoV-2 IgG titres were positive (≥ 10 AU/L) in 108 individuals, negative (< 5 AU/L) in 4411 individuals, and 35 subjects showed borderline results (5–10 AU/mL) (S5 Fig).

To determine the sensitivity and specificity of the screening assay, confirmatory testing was performed in all sera that tested positive for IgM or IgG, all sera with IgG values between 5 and 10 AU/mL, and all sera from SARS-CoV-2 PCR-positive individuals. For confirmation, the total antibodies against SARS-CoV-2 N protein were determined using an electrochemiluminescent immunoassay (ECLIA) on a Cobas e411 analyser (Roche Diagnostics, Mannheim, Germany). In all samples with incongruent results, IgG antibodies against SARS-CoV-2 S1 protein were determined using an enzyme-linked immunosorbent assay (ELISA) (Euroimmun, Luebeck, Germany), while immunoblot was used to differentiate antibodies against N, S1, and the receptor binding domain (RBD) of SARS-CoV-2 from those against seasonal coronaviruses (Mikrogen, Neuried, Germany).

Tests were considered correct if the presence of antibodies was confirmed by at least one more independent assay. Of the 108 sera that tested positive in the Yhlo screening assay, 93 also tested positive in the Roche IgG assay, eight were confirmed by immunoblotting (S1 and S4 Tables). In one individual the screening result was considered specific due to high antibody IgG titre and concomitant IgM positivity although no confirmatory testing could be performed (S1 and S4 Tables, Sample-ID 18). In another individual testing only IgG positive, the serum amount was insufficient for confirmatory testing (S3 Table, Sample-ID 114). Five IgG test results were considered false positive because screening IgG results were not confirmed by any of the other assays (S3 Table). This resulted in a specificity of 99.89% for the IgG assay (4441/4446; S5 Table).

IgM antibodies were screened in all subjects until May 4 (n=1620). Six subjects lacking prevalence of anti-SARS-CoV-2 IgG antibodies tested positive for IgM (6/1620). As this could not be confirmed by the Roche ECLIA detecting IgM and IgG antibodies, these samples were considered false positive (S4 Table). If the Roche assay would have a 100% sensitivity, this would result in a specificity of 99.63% for the IgM assay. Due to the lack of a third assay, this, however, has to be considered preliminary.

To determine the sensitivity of the Yhlo IgG screening assay, 35 samples with detectable values between 5 and 10 AU/mL, i.e. below the recommended cut-off of the assay, were retested with both the Roche and the Euroimmun assay. Four samples tested positive in the Roche and Euroimmun assays, and were therefore considered false negative in the Yhlo screening assay (S1 and S4 Tables). This allowed us to estimate the overall sensitivity of the IgG assay at 96.30% (104/108, S5 Table).

For estimation of seroprevalence, individuals with at least two positive antibody tests (n=106) as well as two individuals with positive SARS-CoV-2 PCR tests that seroconverted during follow-up, were considered seropositive (108/4554) resulting in a seroprevalence of 2•4%. The IgG antibody levels of seropositive individuals were inversely correlated with the time of testing (rho = -0.22, [95% CI -0.39 to -0.03]) (S5 Fig).

From May 5, subjects were tested for anti-SARS-CoV-2 IgM antibodies if specific anti-SARS-CoV-2 IgG antibodies were detected or typical symptoms of COVID-19 were reported (n=88). Overall, concomitant anti-SARS-CoV-2 IgG and IgM was found in 22 subjects (22/1708), of these nine before May 5 (S2 Fig).

S1 Table: Samples with confirmed positive IgG against SARS-CoV-2

| **Sample  ID** | **SARS-CoV-2 PCR** | **YHLO IgG**  **(AU/mL)** | | **YHLO IgM**  **(AU/mL)** | | **Roche IgG ‡ IgM**  **(COI)** | | **Euroimmun IgG** | **Mikrogen recomLine** |
| --- | --- | --- | --- | --- | --- | --- | --- | --- | --- |
| 1 | •• | POSITIVE | (80.12) | POSITIVE | (19.27) | POSITIVE | (14.42) | •• | •• |
| 2 | •• | POSITIVE | (92.39) | POSITIVE | (17.93) | POSITIVE | (52.30) | •• | •• |
| 3 | •• | POSITIVE | (64.82) | POSITIVE | (31.04) | POSITIVE | (25.83) | •• | •• |
| 4 | § | POSITIVE | (113.83) | NEGATIVE | (1.78) | POSITIVE | (66.12) | •• | •• |
| 5 | §‡ | POSITIVE | (93.54) | NEGATIVE | (2.43) | POSITIVE | (21.33) | •• | •• |
| 6 | § | POSITIVE | (109.62) | NEGATIVE | (3.46) | POSITIVE | (49.23) | •• | •• |
| 7 | •• | POSITIVE | (102.26) | NEGATIVE | (2.36) | POSITIVE | (17.38) | •• | •• |
| 8 | •• | POSITIVE | (33.60) | NEGATIVE | (2.80) | POSITIVE | (9.08) | •• | •• |
| 9 | •• | POSITIVE | (28.28) | NEGATIVE | (2.80) | NEGATIVE | (0.06) | NEGATIVE | POSITIVE |
| 10 | •• | POSITIVE | (38.15) | NEGATIVE | (4.40) | POSITIVE | (13.05) | •• | •• |
| 11 | •• | POSITIVE | (113.58) | NEGATIVE | (3.05) | POSITIVE | (65.42) | •• | •• |
| 12 | § | POSITIVE | (96.21) | NEGATIVE | (1.91) | POSITIVE | (47.08) | •• | •• |
| 13 | § | POSITIVE | (78.86) | NEGATIVE | (2.29) | POSITIVE | (32.81) | •• | •• |
| 14 | ‡ | POSITIVE | (107.45) | NEGATIVE | (0.59) | POSITIVE | (14.49) | •• | •• |
| 15 | •• | POSITIVE | (41.99) | NEGATIVE | (2.59) | POSITIVE | (2.98) | •• | •• |
| 16 | ‡ | POSITIVE | (69.45) | NEGATIVE | (0.60) | NEGATIVE | (0.055) | NEGATIVE | POSITIVE |
| 17 | •• | POSITIVE | (84.41) | NEGATIVE | (5.78) | POSITIVE | (40.43) | •• | •• |
| 18 | •• | POSITIVE | (78.97) | POSITIVE | (34.76) | * | •• | * | •• |
| 19 | §‡ | POSITIVE | (12.45) | NEGATIVE | (0.83) | NEGATIVE | (0.45) | BORDERLINE | POSITIVE |
| 20 | •• | POSITIVE | (86.18) | NEGATIVE | (1.14) | POSITIVE | (19.89) | •• | •• |
| 21 | •• | POSITIVE | (22.12) | NEGATIVE | (0.73) | POSITIVE | (1.81) | •• | •• |
| 22 | •• | POSITIVE | (92.35) | POSITIVE | (19.96) | POSITIVE | (28.59) | •• | •• |
| 23 | •• | POSITIVE | (100.84) | POSITIVE | (11.19) | POSITIVE | (84.33) | •• | •• |
| 24 | •• | POSITIVE | (30.86) | NEGATIVE | (0.48) | NEGATIVE | (0.054) | NEGATIVE | POSITIVE |
| 25 | ‡ | POSITIVE | (95.56) | NEGATIVE | (6.06) | POSITIVE | (27.94) | •• | •• |
| 26 | •• | POSITIVE | (91.06) | NEGATIVE | (1.41) | POSITIVE | (48.79) | •• | •• |
| 27 | •• | POSITIVE | (25.61) | NEGATIVE | (1.89) | POSITIVE | (4.55) | •• | •• |
| 28 | § | POSITIVE | (35.34) | NEGATIVE | (1.67) | POSITIVE | (7.32) | •• | •• |
| 29 | •• | POSITIVE | (72.10) | POSITIVE | (22.39) | POSITIVE | (75.58) | •• | •• |
| 30 | § | POSITIVE | (97.45) | NEGATIVE | (4.91) | POSITIVE | (65.48) | •• | •• |
| 31 | § | POSITIVE | (59.51) | POSITIVE | (10.26) | POSITIVE | (79.13) | •• | •• |
| 32 | •• | POSITIVE | (42.14) | POSITIVE | (17.68) | POSITIVE | (14.92) | •• | •• |
| 33 | § | POSITIVE | (49.91) | NEGATIVE | (1.19) | POSITIVE | (27.04) | •• | •• |
| 34 | §‡ | POSITIVE | (97.02) | NEGATIVE | (10.00) | POSITIVE | (99.14) | •• | •• |
| 35 | §‡ | POSITIVE | (91.68) | NEGATIVE | (1.89) | POSITIVE | (14.13) | •• | •• |
| 36 | •• | POSITIVE | (35.20) | NEGATIVE | (1.98) | POSITIVE | (2.63) | •• | •• |
| 37 | § | POSITIVE | (55.52) | NEGATIVE | (0.53) | POSITIVE | (10.45) | •• | •• |
| 38 | •• | POSITIVE | (27.81) | NEGATIVE | (0.48) | POSITIVE | (14.44) | •• | •• |
| 39 | •• | POSITIVE | (98.57) | NEGATIVE | (4.07) | POSITIVE | (70.04) | •• | •• |
| 40 | •• | POSITIVE | (86.47) | NEGATIVE | (9.97) | POSITIVE | (89.22) | •• | •• |
| 41 | •• | POSITIVE | (12.51) | NEGATIVE | (1.06) | POSITIVE | (2.87) | •• | •• |
| **Sample ID** | **SARS-CoV-2 PCR** | **YHLO IgG**  **(AU/mL)** | | **YHLO IgM**  **(AU/mL)** | | **Roche IgG ‡ IgM**  **(COI)** | | **Euroimmun IgG** | **Mikrogen recomLine** |
| 42 | •• | POSITIVE | (45.40) | NEGATIVE | (0.79) | POSITIVE | (21.43) | •• | •• |
| 43 | §‡ | POSITIVE | (40.13) | NEGATIVE | (1.65) | POSITIVE | (25.15) | •• | •• |
| 44 | §‡ | POSITIVE | (35.65) | NEGATIVE | (9.91) | POSITIVE | (51.67) | •• | •• |
| 45 | §‡ | POSITIVE | (91.08) | NEGATIVE | (0.43) | POSITIVE | (89.17) | •• | •• |
| 46 | •• | POSITIVE | (45.18) | NEGATIVE | (1.76) | POSITIVE | (30.29) | •• | •• |
| 47 | §‡ | POSITIVE | (113.23) | NEGATIVE | (2.61) | POSITIVE | (40.58) | •• | •• |
| 48 | •• | POSITIVE | (73.79) | NEGATIVE | (0.67) | POSITIVE | (43.39) | •• | •• |
| 49 | •• | POSITIVE | (93.50) | POSITIVE | (10.14) | POSITIVE | (27.46) | •• | •• |
| 50 | §‡ | POSITIVE | (84.28) | NEGATIVE | (0.68) | POSITIVE | (50.96) | •• | •• |
| 51 | §‡ | POSITIVE | (96.75) | NEGATIVE | (4.80) | POSITIVE | (100.30) | •• | •• |
| 52 | •• | POSITIVE | (36.27) | NEGATIVE | (0.85) | POSITIVE | (2.72) | •• | •• |
| 53 | •• | POSITIVE | (10.52) | NEGATIVE | (0.37) | NEGATIVE | (0.061) | NEGATIVE | POSITIVE |
| 54 | § | POSITIVE | (49.49) | NEGATIVE | (0.94) | POSITIVE | (39.16) | •• | •• |
| 55 | •• | POSITIVE | (50.98) | NEGATIVE | (1.54) | POSITIVE | (78.15) | •• | •• |
| 56 | §‡ | POSITIVE | (83.97) | NEGATIVE | (5.40) | POSITIVE | (45.04) | •• | •• |
| 57 | ‡ | POSITIVE | (32.83) | NEGATIVE | (0.24) | POSITIVE | (19.70) | •• | •• |
| 58 | § | POSITIVE | (72.67) | NEGATIVE | (3.55) | POSITIVE | (11.77) | •• | •• |
| 59 | •• | POSITIVE | (60.13) | NEGATIVE | (2.74) | NEGATIVE | (0.055) | NEGATIVE | POSITIVE |
| 60 | •• | POSITIVE | (66.95) | NEGATIVE | (2.27) | POSITIVE | (39.01) | •• | •• |
| 61 | ‡ | POSITIVE | (47.80) | NEGATIVE | (1.08) | POSITIVE | (19.42) | •• | •• |
| 62 | •• | POSITIVE | (82.89) | NEGATIVE | (4.19) | POSITIVE | (60.83) | •• | •• |
| 63 | § | POSITIVE | (23.44) | NEGATIVE | (0.42) | POSITIVE | (14.57) | •• | •• |
| 64 | •• | POSITIVE | (20.75) | NEGATIVE | (1.49) | POSITIVE | (21.14) | •• | •• |
| 65 | •• | POSITIVE | (45.24) | NEGATIVE | (0.72) | POSITIVE | (53.24) | •• | •• |
| 66 | •• | POSITIVE | (33.84) | NEGATIVE | (0.29) | POSITIVE | (17.67) | •• | •• |
| 67 | •• | POSITIVE | (14.69) | NEGATIVE | (0.49) | POSITIVE | (10.88) | •• | •• |
| 68 | ‡ | POSITIVE | (54.55) | POSITIVE | (22.09) | POSITIVE | (62.24) | •• | •• |
| 69 | ‡ | POSITIVE | (46.11) | NEGATIVE | (0.95) | POSITIVE | (36.16) | •• | •• |
| 70 | ‡ | POSITIVE | (10.01) | NEGATIVE | (1.34) | POSITIVE | (9.90) | •• | •• |
| 71 | •• | POSITIVE | (52.17) | NEGATIVE | (0.62) | POSITIVE | (32.00) | •• | •• |
| 72 | •• | POSITIVE | (13.79) | NEGATIVE | (0.73) | POSITIVE | (8.11) | •• | •• |
| 73 | •• | POSITIVE | (69.38) | POSITIVE | (29.99) | POSITIVE | (91.78) | •• | •• |
| 74 | •• | POSITIVE | (68.40) | NEGATIVE | (4.42) | POSITIVE | (90.23) | •• | •• |
| 75 | •• | POSITIVE | (27.70) | NEGATIVE | (0.40) | POSITIVE | (6.11) | •• | •• |
| 76 | •• | POSITIVE | (82.56) | POSITIVE | (19.32) | POSITIVE | (102.00) | •• | •• |
| 77 | •• | POSITIVE | (58.50) | NEGATIVE | (0.41) | POSITIVE | (84.81) | •• | •• |
| 78 | §‡ | POSITIVE | (74.31) | NEGATIVE | (0.78) | POSITIVE | (68.42) | •• | •• |
| 79 | •• | POSITIVE | (69.18) | NEGATIVE | (0.50) | POSITIVE | (81.05) | •• | •• |
| 80 | •• | POSITIVE | (35.63) | NEGATIVE | (1.11) | POSITIVE | (62.51) | •• | •• |
| 81 | ‡ | POSITIVE | (64.48) | NEGATIVE | (0.80) | POSITIVE | (56.41) | •• | •• |
| 82 | •• | POSITIVE | (82.53) | POSITIVE | (22.54) | POSITIVE | (81.01) | •• | •• |
| 83 | •• | POSITIVE | (10.52) | NEGATIVE | (0.48) | NEGATIVE | (0.053) | NEGATIVE | POSITIVE |
| **Sample ID** | **SARS-CoV-2 PCR** | **YHLO IgG**  **(AU/mL)** | | **YHLO IgM**  **(AU/mL)** | | **Roche IgG ‡ IgM**  **(COI)** | | **Euroimmun  IgG** | **Mikrogen recomLine** |
| 84 | •• | POSITIVE | (68.97) | NEGATIVE | (2.00) | POSITIVE | (51.00) | •• | •• |
| 85 | •• | POSITIVE | (80.39) | NEGATIVE | (5.54) | POSITIVE | (94.25) | •• | •• |
| 86 | ‡ | POSITIVE | (52.70) | POSITIVE | (30.33) | POSITIVE | (40.29) | •• | •• |
| 87 | •• | POSITIVE | (35.43) | NEGATIVE | (0.36) | POSITIVE | (16.54) | •• | •• |
| 88 | § | POSITIVE | (27.04) | NEGATIVE | (5.83) | POSITIVE | (1.15) | •• | •• |
| 89 | •• | POSITIVE | (75.74) | NEGATIVE | (2.60) | POSITIVE | (85.36) | •• | •• |
| 90 | •• | POSITIVE | (57.12) | NEGATIVE | (2.73) | NEGATIVE | (0.055) | NEGATIVE | POSITIVE |
| 91 | •• | POSITIVE | (26.50) | NEGATIVE | (0.45) | POSITIVE | (52.46) | •• | •• |
| 92 | •• | POSITIVE | (63.67) | NEGATIVE | (0.46) | POSITIVE | (31.93) | •• | •• |
| 93 | •• | POSITIVE | (56.33) | NEGATIVE | (0.67) | POSITIVE | (100.70) | •• | •• |
| 94 | •• | POSITIVE | (85.75) | NEGATIVE | (0.66) | POSITIVE | (112.80) | •• | •• |
| 95 | •• | POSITIVE | (41.98) | NEGATIVE | (0.63) | POSITIVE | (87.69) | •• | •• |
| 96 | •• | POSITIVE | (14.15) | NEGATIVE | (0.97) | POSITIVE | (11.00) | •• | •• |
| 97 | § | POSITIVE | (83.09) | NEGATIVE | (1.92) | POSITIVE | (112.00) | •• | •• |
| 98 | •• | POSITIVE | (43.75) | NEGATIVE | (0.63) | POSITIVE | (77.74) | •• | •• |
| 99 | ‡ | POSITIVE | (54.46) | NEGATIVE | (22.17) | POSITIVE | (73.22) | •• | •• |
| 100 | §‡ | POSITIVE | (86.86) | NEGATIVE | (1.45) | POSITIVE | (70.92) | •• | •• |
| 101 | •• | POSITIVE | (15.87) | NEGATIVE | (1.08) | POSITIVE | (3.05) | •• | •• |
| 102 | §‡ | POSITIVE | (63.04) | NEGATIVE | (5.02) | POSITIVE | (120.80) | •• | •• |
| 103 | §‡ | NEGATIVE | (6.55) | •• | •• | POSITIVE | (1.76) | POSITIVE | •• |
| 104 | ‡ | NEGATIVE | (5.26) | •• | •• | POSITIVE | (2.22) | POSITIVE | •• |
| 105 | •• | NEGATIVE | (8.81) | •• | •• | POSITIVE | (5.85) | POSITIVE | •• |
| 106 | ‡ | NEGATIVE | (6.64) | •• | •• | POSITIVE | (1.75) | POSITIVE | •• |
| 107† | § | POSITIVE† | (37.99) | POSITIVE† | (20.47) | •• | •• | •• | •• |
| 108† | § | POSITIVE† | (45.70) | NEGATIVE | (1.14) | •• | •• | •• | •• |

* No material for further tests available, † positive at follow-up visit, COI: Cut-off index, ‡ positive SARS-CoV-2 PCR extern, § positive SARS-CoV-2 PCR in-house, •• not available

S2 Table: Summary of confirmatory assays

| **YHLO IgG**  **(AU/ml)** | **YHLO IgM**  **(AU/ml)** | **Roche IgG ‡ IgM**  **(COI)** | **Euroimmun IgG**  **(ratio)** | **Mikrogen recomLine**  **immunoblot** | **Final result** | **No. of subjects** |
| --- | --- | --- | --- | --- | --- | --- |
| POSITIVE | •• | POSITIVE | •• | •• | POSITIVE | 93 |
| POSITIVE | •• | NEGATIVE | NEGATIVE | POSITIVE | POSITIVE | 8 |
| POSITIVE | NEGATIVE | NEGATIVE | NEGATIVE/BORDERLINE | NEGATIVE | NEGATIVE | 5 |
| POSITIVE | POSITIVE | * | * | * | POSITIVE | 1 |
| POSITIVE | NEGATIVE | * | * | NEGATIVE | Excluded from calculation of specificity | 1 |
| BORDERLINE | •• | POSITIVE | POSITIVE | •• | POSITIVE | 4 |
| POSITIVE† | •• | •• | •• | •• | POSITIVE | 2^§^ |

† Initially negative, but positive at follow-up visit, * no material for further tests available, ^§^ in-house SARS-CoV-2 PCR positive, COI: Cut-off index

S3 Table: Samples with positive IgG against SARS-CoV-2 that could not be confirmed

| **Sample ID** | **YHLO IgG**  **(AU/mL)** | | **YHLO IgM**  **(AU/mL)** | | **Roche IgG ‡ IgM**  **(COI)** | | **Euroimmun IgG** | **Mikrogen recomLine** |
| --- | --- | --- | --- | --- | --- | --- | --- | --- |
| 109 | POSITIVE | (26.78) | NEGATIVE | (0.19) | NEGATIVE | (0.079) | NEGATIVE | NEGATIVE |
| 110 | POSITIVE | (23.75) | NEGATIVE | (1.17) | NEGATIVE | (0.055) | BORDERLINE | NEGATIVE |
| 111 | POSITIVE | (10.14) | NEGATIVE | (0.82) | NEGATIVE | (0.126) | NEGATIVE | NEGATIVE |
| 112 | POSITIVE | (21.46) | NEGATIVE | (0.44) | NEGATIVE | (0.102) | NEGATIVE | NEGATIVE |
| 113 | POSITIVE | (12.13) | NEGATIVE | (0.50) | NEGATIVE | (0.055) | NEGATIVE | NEGATIVE |
| 114 | POSITIVE | (11.75) | NEGATIVE | (0.28) | * | •• | * | •• |

* No material for further tests available

S4 Table: Samples with positive IgM against SARS-CoV-2 that could not be confirmed

| **Sample ID** | **YHLO IgG**  **(AU/mL)** | | **YHLO IgM**  **(AU/mL)** | | **Roche IgG ‡ IgM**  **(COI)** | |
| --- | --- | --- | --- | --- | --- | --- |
| 115 | NEGATIVE | (0.98) | POSITIVE | (11.51) | NEGATIVE | (0.054) |
| 116 | NEGATIVE | (0.41) | POSITIVE | (13.71) | NEGATIVE | (0.055) |
| 117 | NEGATIVE | (2.90) | POSITIVE | (13.41) | NEGATIVE | (0.055) |
| 118 | NEGATIVE | (0.28) | POSITIVE | (12.23) | NEGATIVE | (0.056) |
| 119 | NEGATIVE | (0.45) | POSITIVE | (10.27) | NEGATIVE | (0.056) |
| 120 | NEGATIVE | (0.14) | POSITIVE | (265.82) | NEGATIVE | (0.056) |

S5 Table: Calculation of specificity and sensitivity

|  | **≥ 2 Positive confirmatory tests or concomitant IgM** | **< 2 Positive confirmatory tests** | **Total** |
| --- | --- | --- | --- |
| **Screening assay positive** | True positive  n =104 | False positive  n = 5 | n = 109† |
| **Screening assay negative** | False negative  n = 4 | True negative  n = 4441 | n = 4445 |
| **Total** | n = 108 | n = 4446 | 4554 |

†Serum material was insufficient for confirmatory testing in one seropositive subject, two subjects seroconverted at follow-up visit.

S1 Figure

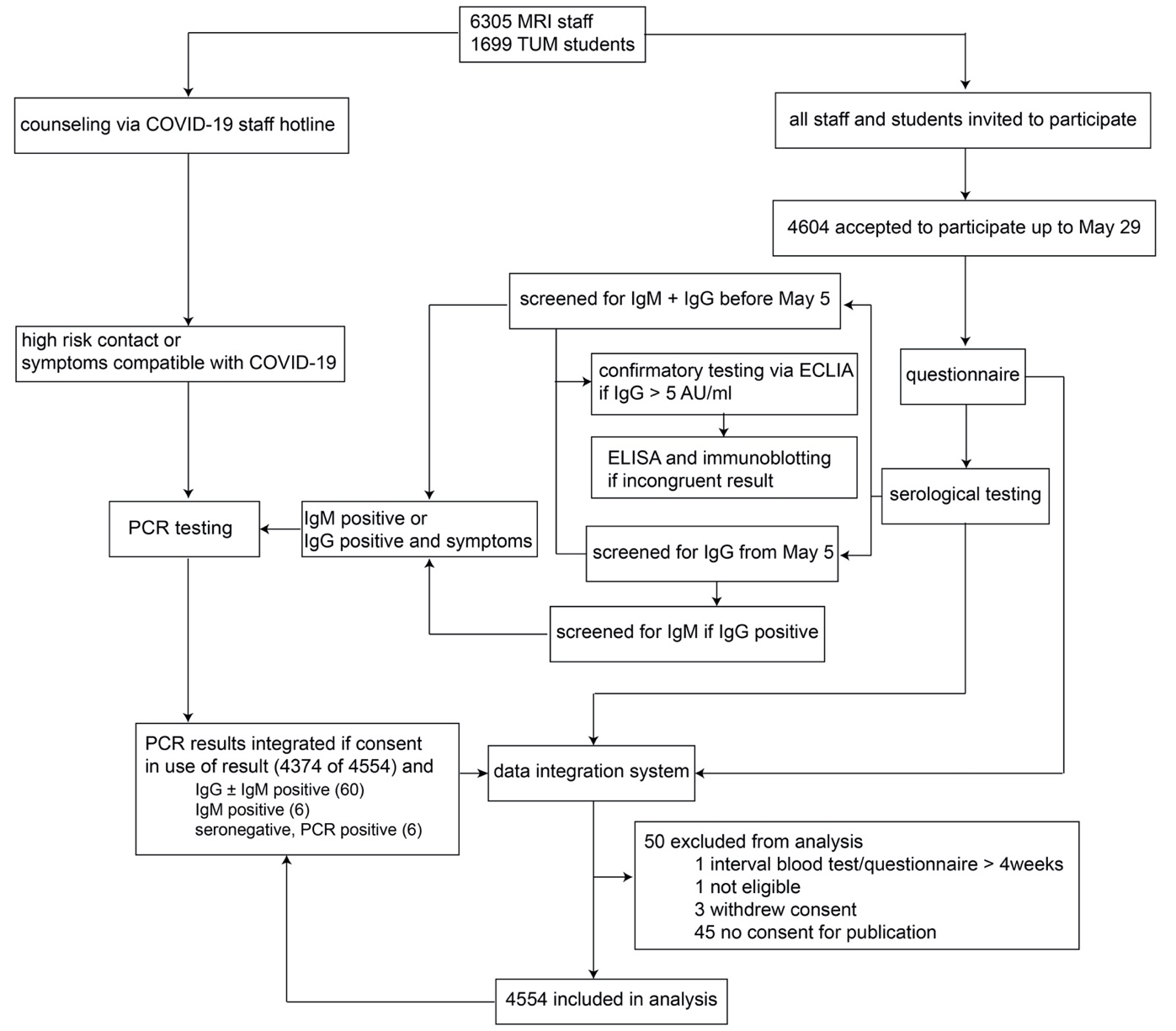

**S1 Fig: Study flow chart illustrating the testing algorithm and included results**

CLIA: Chemiluminescent immunoassay, ECLIA: Electrochemiluminescent immunoassay, ELISA: Enzyme-linked immunosorbent assay, MRI: Munich rechts der Isar Hospital, PCR: Polymerase chain reaction, TUM: Technical University Munich

S2 Figure

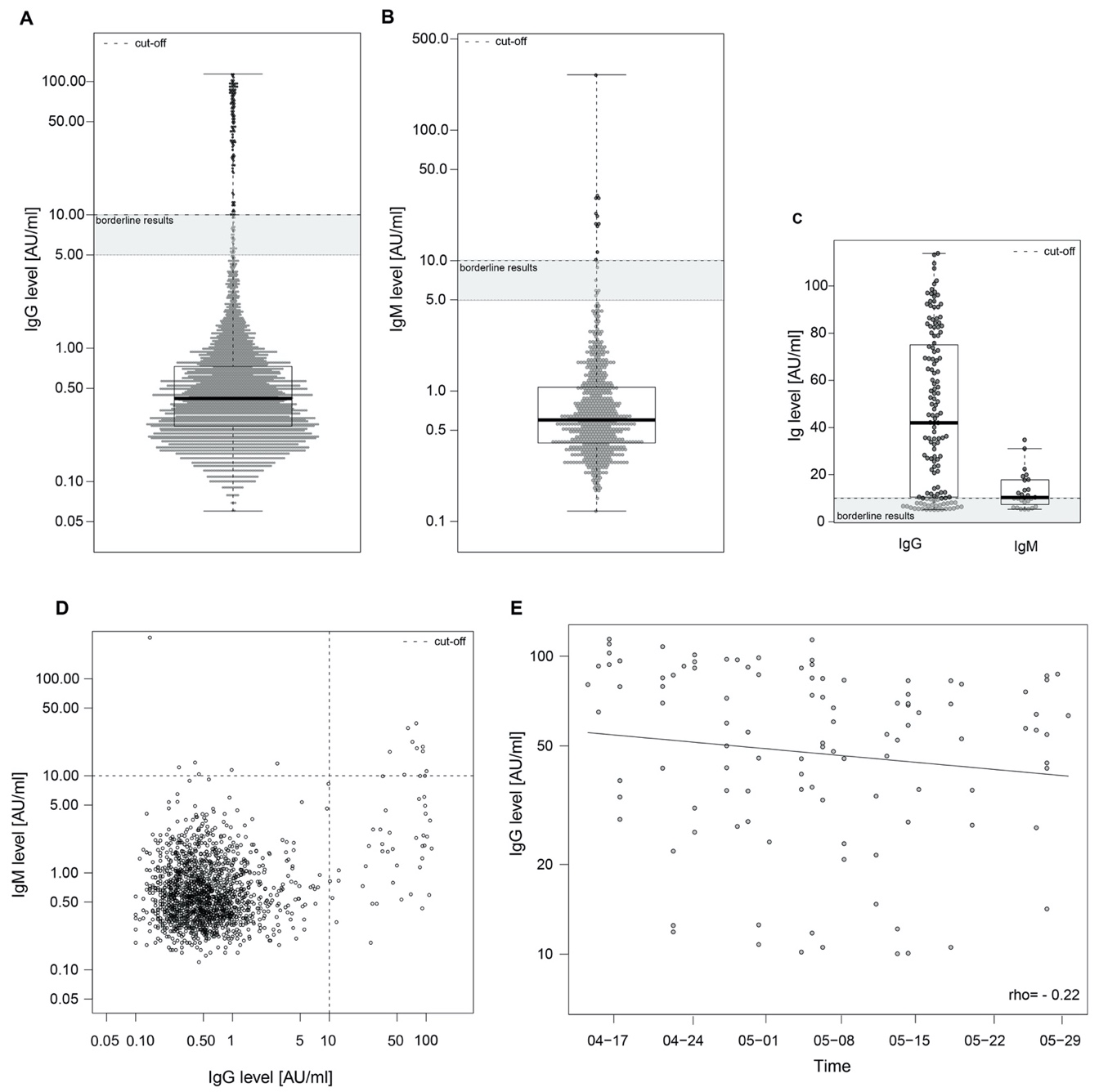

**S2 Fig: Anti-SARS-CoV-2 IgG and IgM levels**
Anti-SARS-CoV-2 IgG and IgM antibodies were detected using a paramagnetic particle chemiluminescent immunoassay (Shenzhen Yloh Biotech; Shenzhen, China). The cut-off was defined as ≥ 10 AU/mL per assay instruction, and is indicated by a dashed line. Boxplots show medians (thick middle line), as well as first and third quartiles (box boundaries), while the whiskers indicate ranges. IgG antibodies were measured in all participants (n = 4554) (A). The IgM levels of all participants tested up to May 4, 2020 (n = 1620) are depicted in (B). Thereafter, IgM was only tested in cases with positive IgG results (n = 88, data not shown). All positive IgG and IgM results (up to May 4), as well as borderline results (> 5 AU/L and < 10) are depicted in (C). (D) Shows the correlation of IgG and IgM levels of all participants tested for both immunoglobulins (n=1620 = 1708). (E) IgG levels detected in seropositive individuals are plotted per study day. Spearman’s rank correlation coefficient was used to evaluate the association between the time point of IgG testing and the IgG titre level.

S3 Figure

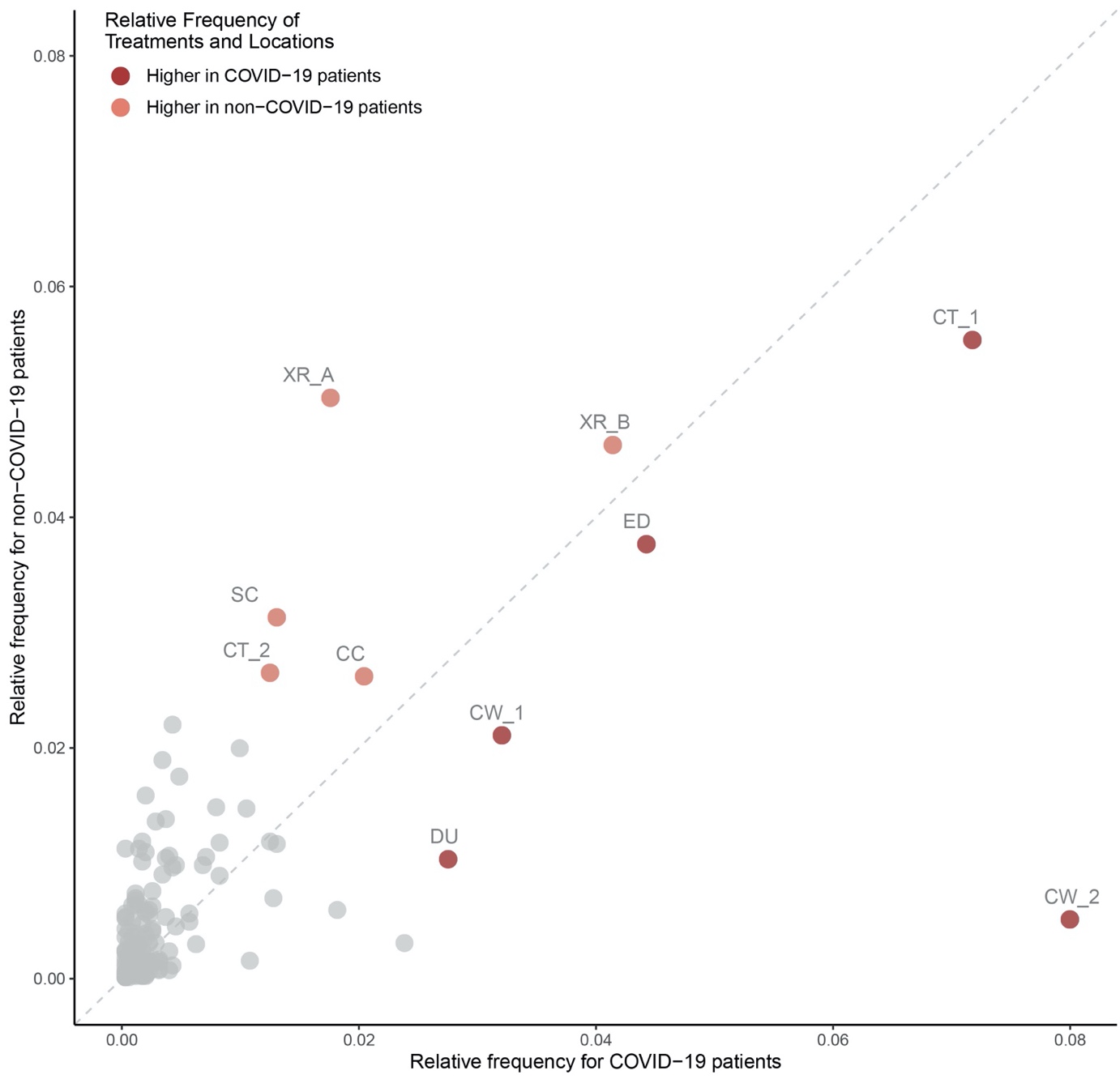

**S3 Fig: Relative frequency of requested diagnostics, therapies, and spatial information between patients diagnosed with COVID-19 and non-COVID-19 pneumonia from December 1, 2019 to June 10, 2020 normalized by each patient group**

Diagram demonstrating that the diagnostic and therapeutic facilities for patients with COVID-19 and non-COVID-19 pneumonia were used differentially by the two patient groups, further limiting the possibilities of infection. CC: Cardiovascular clinic, CT_1 and CT_2: Spatially distinct CT scanners, CW_1: COVID-19 admission ward, CW_2: COVID-19 ward, DU: dialysis unit, ED: Emergency department, SC: Social counselling, XR_A and XR_B: Spatially distinct X-ray units.

S4 Figure

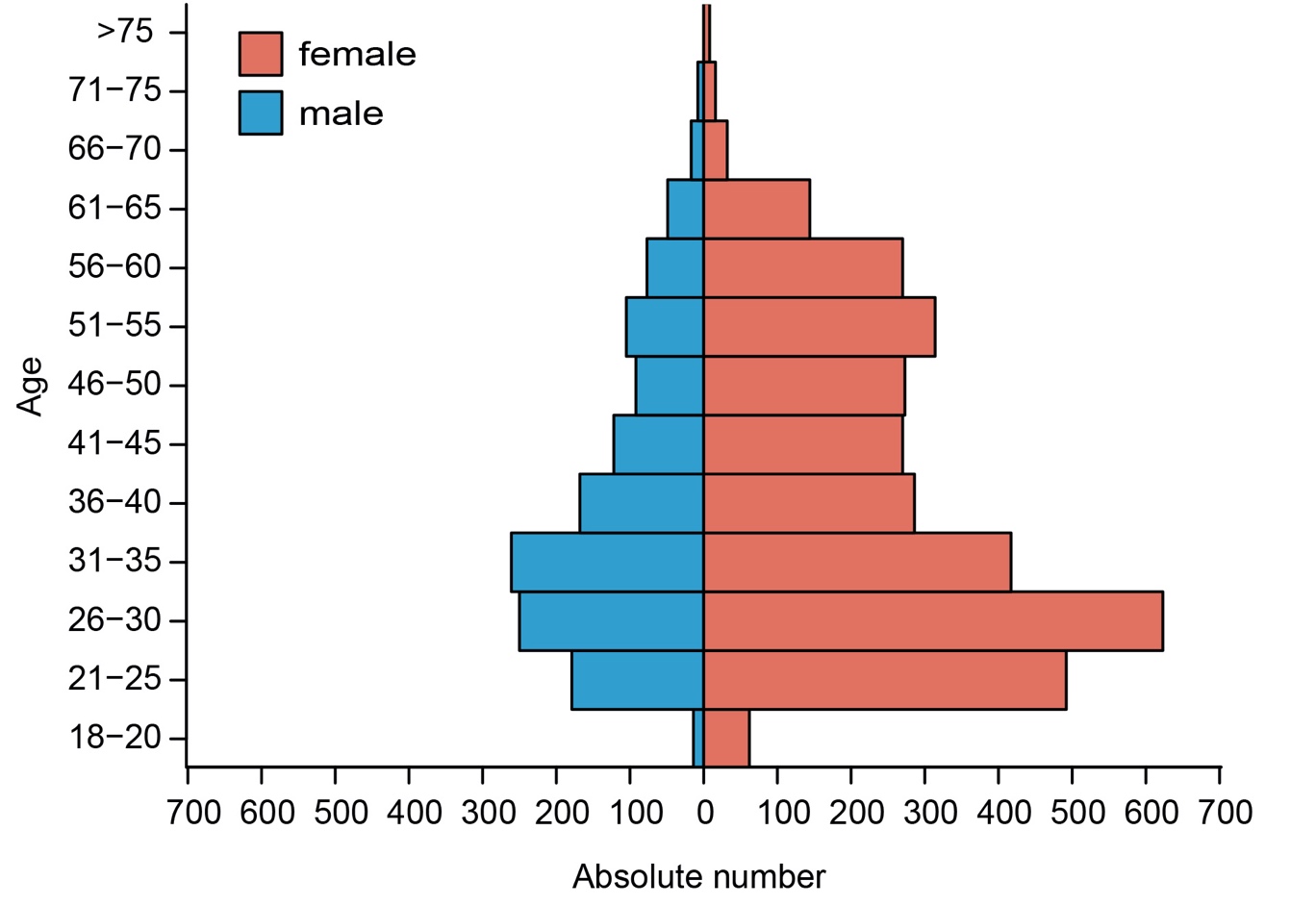

**S4 Fig: Age and sex distribution of the study participants**
The population pyramid depicts the age and sex distribution of the subjects.

S5 Figure

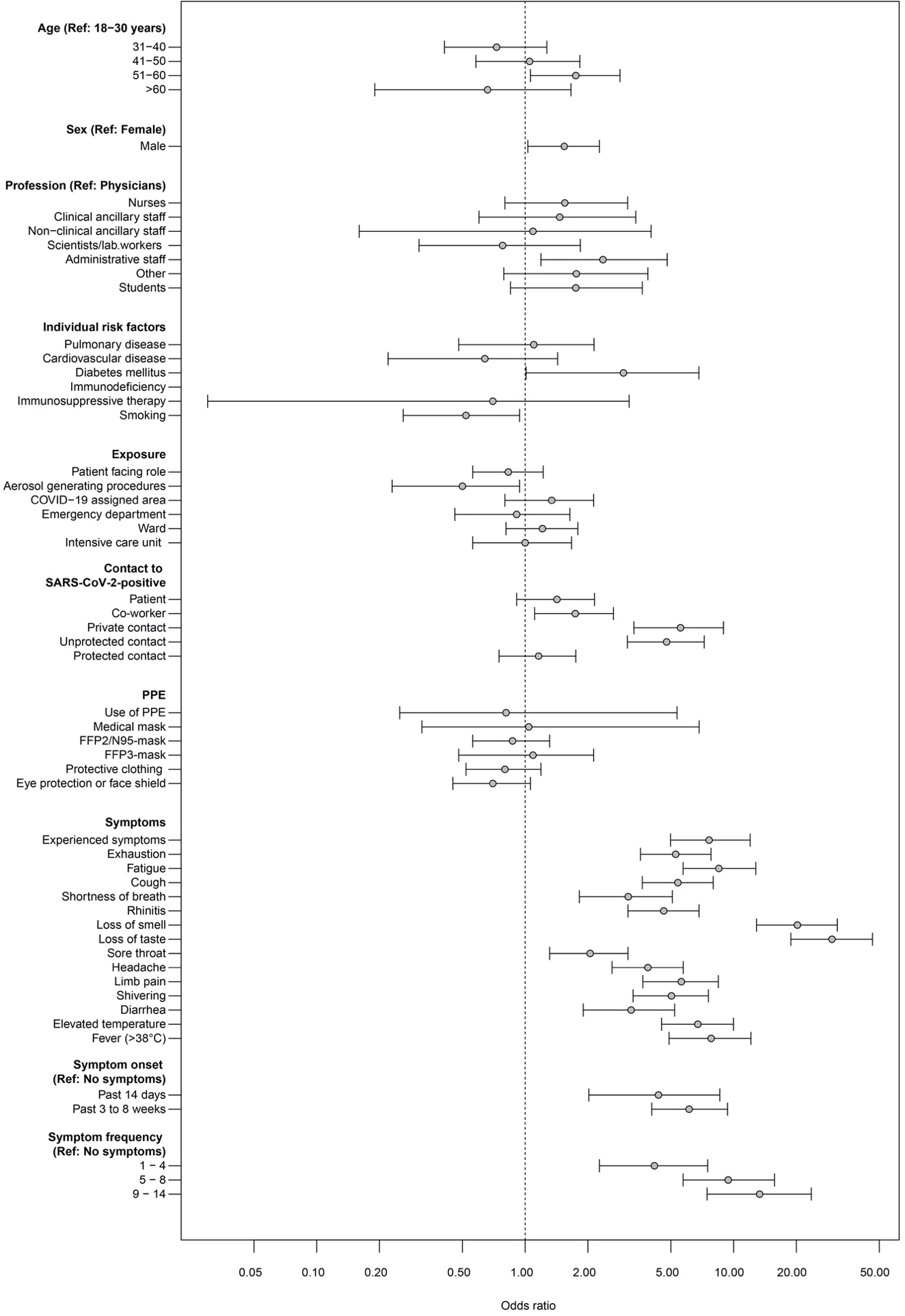

**S5 Fig: Graphic representation of all odds ratios for seropositivity to SARS-CoV-2 IgG represented in S1–S3 Tables**

Odds ratios with exact 95% confidence intervals (mid-p intervals) are presented. FFP: Filtering face piece, FFP 2: Use of masks with 94% or ≥ 95% filter capacity for particles > 0.6 µm, FFP3: Use of masks with 99% filter capacity for particles > 0.6 µm, Lab: Laboratory, Ref: Reference, PPE: Personal protective equipment.

S6 Figure

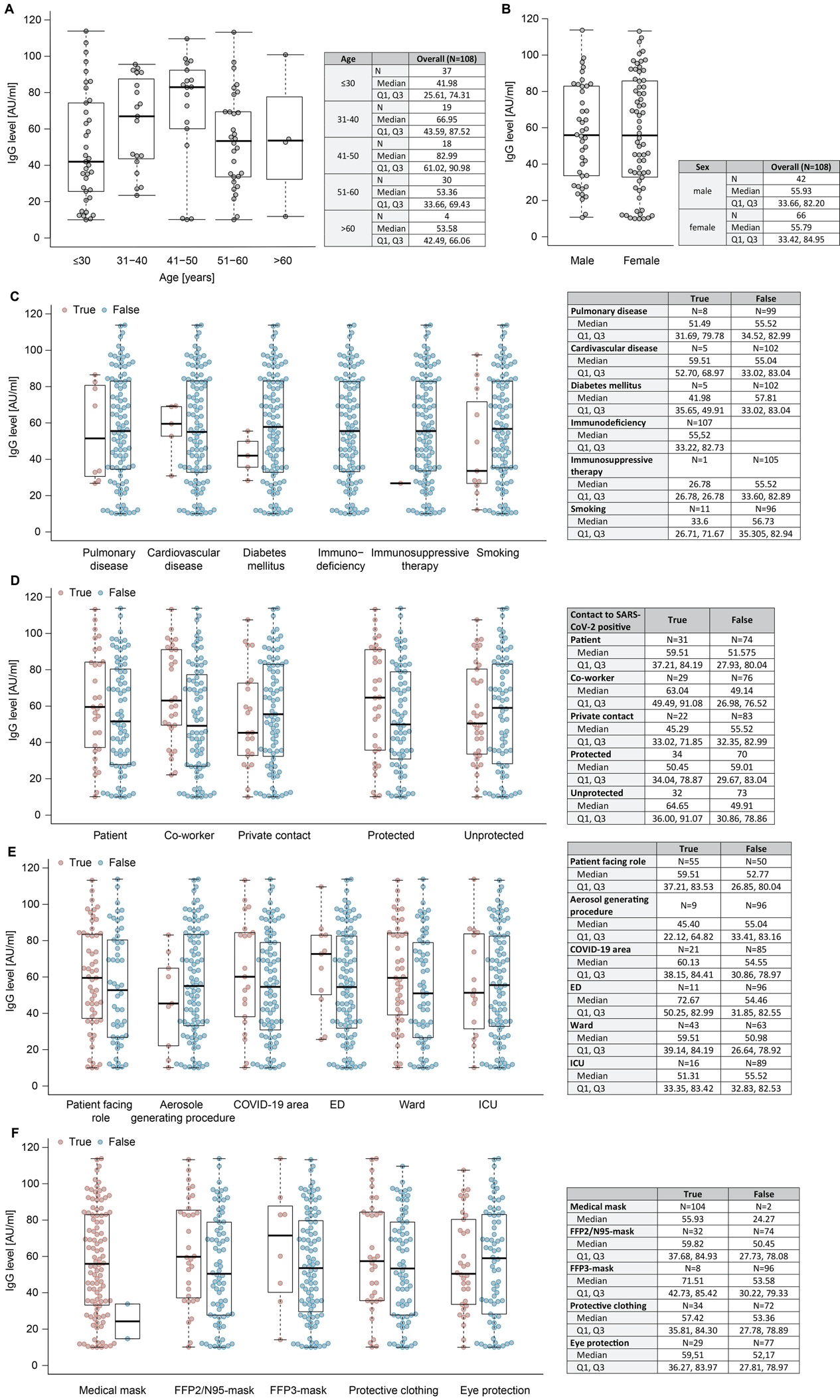

**S6 Fig: Distribution of anti-SARS-CoV-2 IgG levels stratified for personal/occupational risk factors**

IgG levels were compared in seropositive staff of different age (A), sex (B), and with different comorbidities or smoking status (C), reported COVID-19 contact (D), as well as occupational exposures (E) and use of distinct personal protective equipment. Boxplots show medians (thick middle line), as well as the first and third quartiles (box boundaries), while the whiskers indicate ranges. Medians and quartiles are depicted in the adjacent table. ED: Emergency department, FFP: Filtering face piece, ICU: Intensive care unit.

S7 Figure

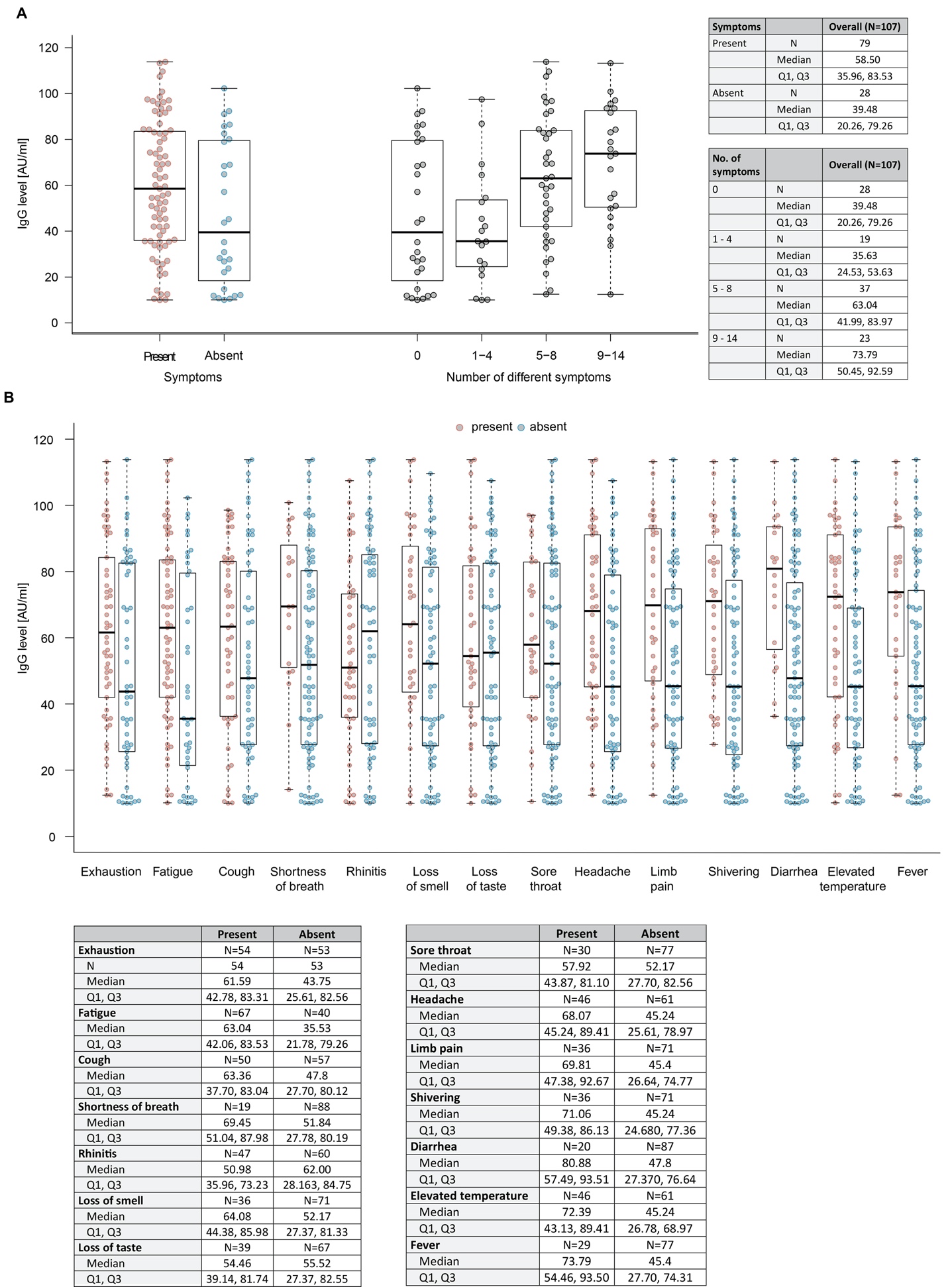

**S7 Fig: Anti-SARS-CoV-2 IgG levels and symptoms**

Distribution of antibodies stratified for symptom frequency (A) and character (B). Boxplots show medians (thick middle line), as well as first and third quartiles (box boundaries). The values are annotated in the adjacent table. The whiskers indicate ranges. Q: Quartile.

S8 Figure

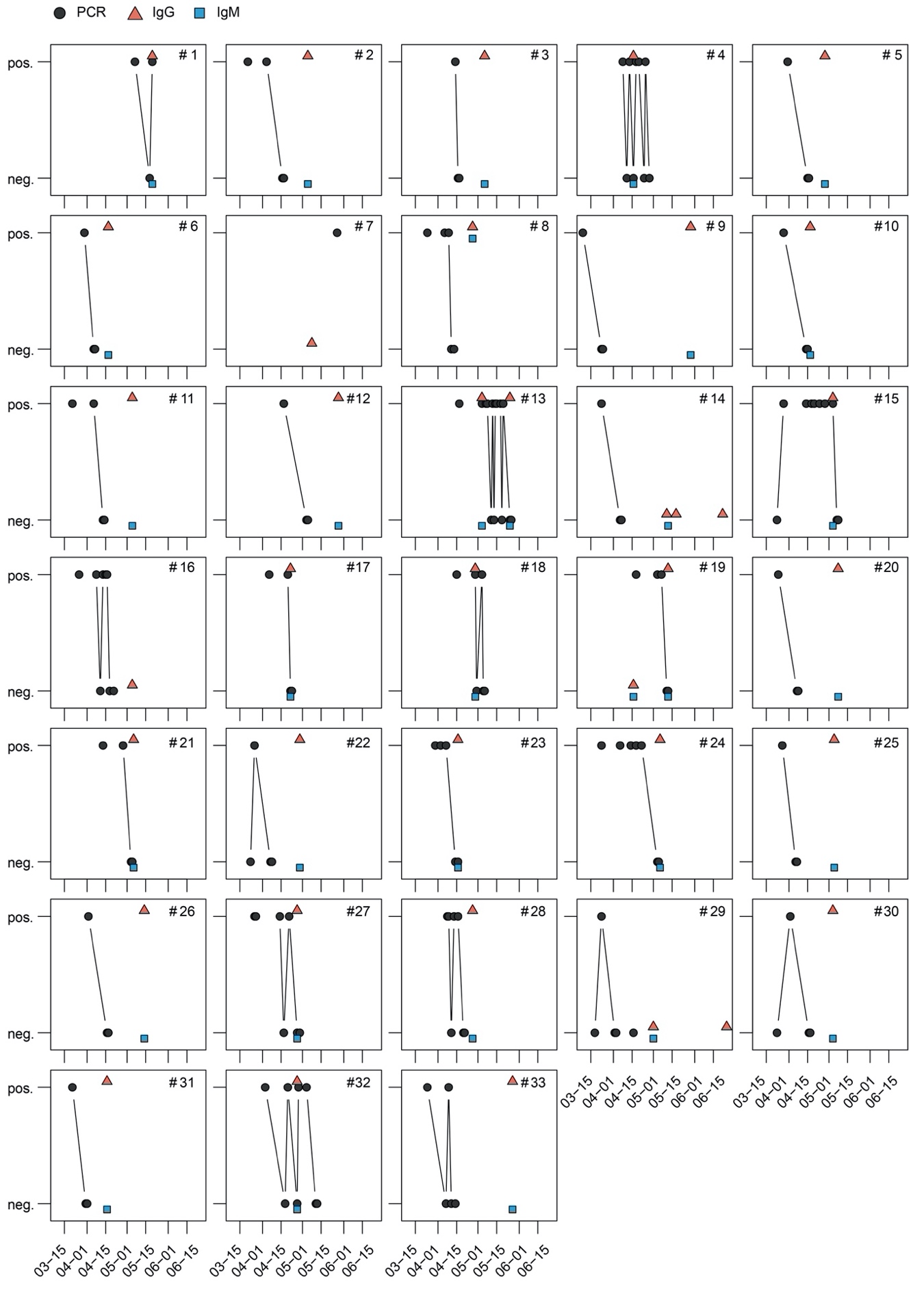

**S8 Fig: Time course of anti-SARS-CoV-2 antibody and PCR test results in 33 employees**

PCR test results were available for 33 employees (numbered consecutively) who had tested positive for SARS-CoV-2 via PCR at least once. Plots show all anti-SARS-CoV-2 IgG and IgM, as well as PCR tests performed before June 15, 2020, in these subjects. Five subjects (No.’s 7, 14, 16, 19, and 29) tested negative for IgG antibodies at the time of the serosurvey. Seroconversion could be detected in No. 19 on follow-up assessment. Pos: Positive, Neg: Negative.

S1 Video

**Time lapse of the relative frequencies of all trajectories for COVID-19 cases**

Frequencies in distinct hospital locations were normalized by all timeframes between February 1, 2020 and May 29, 2020. Some patients who later tested positive for SARS-CoV2 were already in the hospital prior to March 6, 2020, and are therefore visualized starting February 1.

S2 Video

**Time lapse of the relative frequencies of trajectories available for SARS-CoV-2-positive staff working on campus**

Frequencies were mapped for 14 days on their last location before staff either tested positive or were sent to quarantine. Results were normalized by all timeframes between March 22, 2020 to May 29, 2020.

S3 Video

**Time lapse of the relative frequencies for the difference in trajectories available for COVID-19 cases and for SARS-CoV-2-positive staff**

Frequencies in distinct hospital locations were normalised by all timeframes between February 1, 2020 and May 29, 2020.

For the purposes of discretion, graphic representation of spatial information is partially distorted.

STROBE Statement—Checklist of items that should be included in reports of ***cross-sectional studies***

|  | Item No | Recommendation | Page No |
| --- | --- | --- | --- |
| **Title and abstract** | 1 | (*a*) Indicate the study’s design with a commonly used term in the title or the abstract | 3 |
|  |  | (*b*) Provide in the abstract an informative and balanced summary of what was done and what was found | 3 |
| Introduction | | | |
| Background/rationale | 2 | Explain the scientific background and rationale for the investigation being reported | 5 |
| Objectives | 3 | State specific objectives, including any prespecified hypotheses | 5 |
| Methods | | | |
| Study design | 4 | Present key elements of study design early in the paper | 7 |
| Setting | 5 | Describe the setting, locations, and relevant dates, including periods of recruitment, exposure, follow-up, and data collection | 7 |
| Participants | 6 | (*a*) Give the eligibility criteria, and the sources and methods of selection of participants | 7 |
| Variables | 7 | Clearly define all outcomes, exposures, predictors, potential confounders, and effect modifiers. Give diagnostic criteria, if applicable | 7 |
| Data sources/ measurement | 8* | For each variable of interest, give sources of data and details of methods of assessment (measurement). Describe comparability of assessment methods if there is more than one group | 7-9 |
| Bias | 9 | Describe any efforts to address potential sources of bias | 17 |
| Study size | 10 | Explain how the study size was arrived at | 7 |
| Quantitative variables | 11 | Explain how quantitative variables were handled in the analyses. If applicable, describe which groupings were chosen and why | 10 |
| Statistical methods | 12 | (*a*) Describe all statistical methods, including those used to control for confounding | 10 |
|  |  | (*b*) Describe any methods used to examine subgroups and interactions | 10 |
|  |  | (*c*) Explain how missing data were addressed | 10 |
|  |  | (*d*) If applicable, describe analytical methods taking account of sampling strategy | N/A |
|  |  | (*e*) Describe any sensitivity analyses | S3 Appendix |
| Results | | | |
| Participants | 13* | (a) Report numbers of individuals at each stage of study—eg numbers potentially eligible, examined for eligibility, confirmed eligible, included in the study, completing follow-up, and analysed | 11 |
|  |  | (b) Give reasons for non-participation at each stage | S1 Fig |
|  |  | (c) Consider use of a flow diagram | S1 Fig |
| Descriptive data | 14* | (a) Give characteristics of study participants (eg demographic, clinical, social) and information on exposures and potential confounders | 11, 12 |
|  |  | (b) Indicate number of participants with missing data for each variable of interest | 23, 24 |
| Outcome data | 15* | Report numbers of outcome events or summary measures | 11 |
| Main results | 16 | (*a*) Give unadjusted estimates and, if applicable, confounder-adjusted estimates and their precision (eg, 95% confidence interval). Make clear which confounders were adjusted for and why they were included | 11, 12 |
|  |  | (*b*) Report category boundaries when continuous variables were categorized |  |
|  |  | (*c*) If relevant, consider translating estimates of relative risk into absolute risk for a meaningful time period |  |
| Other analyses | 17 | Report other analyses done—eg analyses of subgroups and interactions, and sensitivity analyses | 12-14 |
| Discussion | | | |
| Key results | 18 | Summarise key results with reference to study objectives | 14 |
| Limitations | 19 | Discuss limitations of the study, taking into account sources of potential bias or imprecision. Discuss both direction and magnitude of any potential bias | 17 |
| Interpretation | 20 | Give a cautious overall interpretation of results considering objectives, limitations, multiplicity of analyses, results from similar studies, and other relevant evidence | 18 |
| Generalisability | 21 | Discuss the generalisability (external validity) of the study results | 14 |
| Other information | | | |
| Funding | 22 | Give the source of funding and the role of the funders for the present study and, if applicable, for the original study on which the present article is based | Online portal |

*Give information separately for exposed and unexposed groups.

**Note:** An Explanation and Elaboration article discusses each checklist item and gives methodological background and published examples of transparent reporting. The STROBE checklist is best used in conjunction with this article (freely available on the Web sites of PLoS Medicine at http://www.plosmedicine.org/, Annals of Internal Medicine at http://www.annals.org/, and Epidemiology at http://www.epidem.com/). Information on the STROBE Initiative is available at www.strobe-statement.org.
